# Targeting memory consolidation in major depression via pharmacological slow-wave sleep promotion

**DOI:** 10.64898/2026.11.18.26368401

**Authors:** Laura K. Schnider, Boris B. Quednow, Benjamin Stucky, Jannick Mauron, Julia di Floriano, Björn Rasch, Dario A. Dornbierer, Oliver G. Bosch, Erich Seifritz, Hans-Peter Landolt, Francesco Bavato

## Abstract

Slow-wave sleep (SWS) has been implicated in memory consolidation and brain plasticity and proposed as a therapeutic target in major depressive disorder (MDD). However, whether pharmacological promotion of SWS causally improves memory consolidation remains unknown. Gamma-hydroxybutyrate (GHB), a robust enhancer of SWS, represents a promising drug-repurposing candidate to target this pathway. We therefore conducted a randomized, double-blind, placebo-controlled crossover trial in 22 patients with MDD and 20 matched healthy controls receiving single nocturnal doses of GHB, trazodone as an active comparator, or placebo under polysomnographic monitoring (ClinicalTrials.gov: NCT04082806). Declarative, procedural and emotional memory were assessed before and after sleep. GHB strongly increased SWS duration and delta power, whereas trazodone produced smaller changes in sleep-stage composition. Contrary to our hypothesis, neither drug improved memory consolidation across domains. Exploratory analyses revealed weak associations between global sleep-stage measures and memory, alongside selective associations with spindle and spectral features. These findings challenge the assumption that increasing SWS alone is sufficient to improve overnight memory consolidation and suggest that more targeted modulation of sleep oscillatory dynamics may be required.

## Introduction

Sleep plays a central role in memory consolidation, particularly during non-rapid eye movement (NREM) sleep. Converging evidence indicates that newly acquired information is stabilized and integrated during sleep through coordinated neurophysiological processes involving slow-waves, spindles, and their temporal coupling, which facilitate hippocampal-neocortical communication [1] [2] [3]. During slow-wave sleep (SWS), cortical down-states are thought to permit a renormalization of synaptic weights potentiated during prior wakefulness, while down-to-up transitions provide windows in which hippocampal sharp-wave ripples, nested within spindle troughs, reactivate newly encoded traces and drive their neocortical integration [4] [5]. While some experimental studies in healthy individuals showed that SWS manipulation, including pharmacological and stimulation approaches and nap paradigms, can influence memory consolidation [6] [7] [8] [9], overall findings have been mixed and evidence in clinical populations with disturbed memory performance remains scarce [10] [11] [12].

This question is particularly relevant in major depressive disorder (MDD), a mood disorder characterized by sleep disturbances and memory impairments [13]. Patients with MDD often exhibit reduced NREM slow-wave sleep (SWS), altered spindle activity, and deficits in hippocampus-dependent declarative memory consolidation [14] [15]. These abnormalities have been linked to altered synaptic plasticity [16] and may contribute not only to cognitive dysfunction but also to poorer treatment response, as effective psychotherapy relies on the consolidation and integration of newly acquired emotional and cognitive information [17]. Furthermore, a negative memory bias, characterized by preferential consolidation of negative information, has been proposed as a mechanism contributing to symptom persistence and relapse in MDD [18]. Modulating memory consolidation through SWS promotion may therefore represent a therapeutic target for precision interventions.

Despite this evidence, current pharmacological treatments for sleep disturbances in depression remain limited. Commonly used sleep compounds, such as benzodiazepines and z-drugs, often have long half-life times, may impair next-day functioning, and show heterogeneous effects on SWS and memory consolidation [19] [20]. Trazodone, a sedating antidepressant widely prescribed off-label for insomnia in patients with MDD, shows variable effects on SWS [21], and has not been systematically evaluated for its impact on memory consolidation. Gamma-hydroxybutyrate (GHB), administered as sodium oxybate, is an endogenous GABA metabolite and an agonist at the GABA_B_ receptor and at the GHB binding site [22]. We previously showed that a single administration of GHB robustly increases SWS and reduces sleep fragmentation without substantial residual effects on next-morning cognitive performance in both healthy controls (HC) and patients with MDD [23] [24] [25]. However, the effects of GHB on memory consolidation, particularly in MDD, is unclear [7].

Here, we tested whether pharmacological promotion of SWS enhances overnight memory consolidation in patients with MDD. In a within-subject design, we conducted a randomized, double-blind, placebo-and active comparator-controlled crossover trial (NCT04082806) in patients with MDD and matched HC. Following a screening and adaptation night, participants completed three experimental nights separated by one-week washout periods, receiving placebo, GHB, and trazodone in randomized order. Overnight polysomnography was performed in a controlled sleep laboratory, and participants completed a battery of memory tasks assessing declarative, procedural, and emotional memory, with encoding before sleep and recall after awakening. Beyond sleep macrostructure, we quantified slow-wave and spindle characteristics, their temporal coupling, and spectral measures to explore macro-and microstructural sleep predictors of overnight memory consolidation. The within-subject design, together with the inclusion of a matched HC group, allowed us to assess the effects of pharmacologically enhanced SWS on memory consolidation and to compare these effects between clinical and healthy populations. The effects of GHB on sleep macrostructure in individuals with MDD from this trial have been reported previously [25]. We hypothesized that pharmacological enhancement of SWS with GHB would improve overnight memory consolidation, particularly hippocampus-dependent declarative memory, with the largest effects expected in patients with MDD, in whom declarative memory consolidation is frequently impaired. Our aim was to provide mechanistic proof-of-concept for SWS-targeted interventions to enhance sleep-dependent memory consolidation in MDD.

## Methods

### Permission

The study was conducted as a single-center trial at the University of Zurich, Switzerland. It received formal approval from the Swiss Agency for Therapeutic Products (Swissmedic Nr. 2018-01283) and the Ethics Committee of the Canton of Zurich. All participants provided written informed consent in accordance with the Declaration of Helsinki and were compensated monetarily for their participation. The study was preregistered on ClinicalTrials.gov (identifier: NCT04082806).

### Study design

The study followed a randomized, placebo-controlled, double-blind, crossover design, comparing GHB with the clinical alternative trazodone and placebo (see Figure 1). All assessments were conducted in the sleep laboratory at the Institute of Pharmacology and Toxicology, University of Zurich, Switzerland. Two study groups, individuals with MDD and HC, were included to distinguish the pharmacological effects of GHB and trazodone from those modulated by depressive disorder. The study protocol involved five nights, each one including an all-night PSG measurement: a screening night in the frame of a medical examination, an adaptation night to allow acclimatization to the laboratory environment, and three experimental nights. The first experimental night followed directly after the adaptation night. Each of the three experimental nights was separated by a 7-day washout period. On the experimental nights, participants were free to choose their sleep-wake rhythm, with a sleep duration of 7.5h and a bedtime between 22 to 24h. For reasons of simplicity, in the following it is only referred to the 24.00-7.30h sleep rhythm. In addition to the results presented here, the study also evaluated drug effects on toxicological urine markers and clinical effects of GHB on clinical scores and sleep in the MDD study group, both published separately [25] [26].

**Figure 1.**
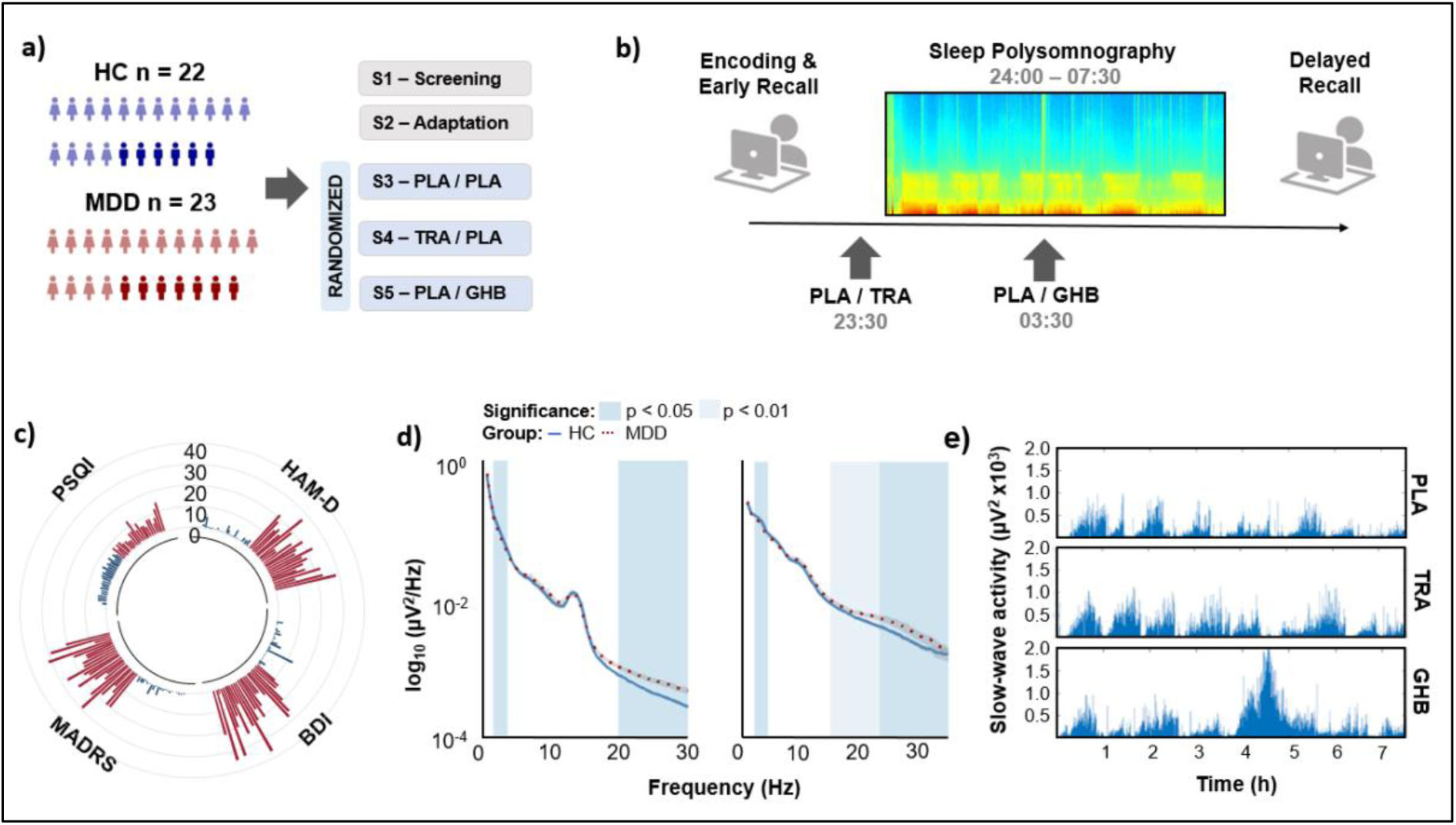
Study overview. **a)** Twenty-two healthy controls (HC, blue) and twenty-three patients with major depressive disorder (MDD, red) were randomized and received at least one treatment. The study design consisted of one screening night (S1), one adaptation night (S2), and three experimental nights (S3–S5) presented in randomized order. **b)** Timeline of an experimental night. Study medications were administered at two time points during the night: trazodone (TRA, 1.5mg/kg) or solid placebo (PLA) at 23:30, and gamma-hydroxybutyrate (GHB, 50mg/kg) or liquid placebo (PLA) at 03:30. Memory task consolidation took place before sleep, and delayed recall occurred after awakening. **c)** Circular histogram of depression and sleep-related scores for healthy controls and patients (PSQI, Pittsburgh Sleep Quality Index; HAM-D, Hamilton Depression Rating Scale; MADRS, Montgomery–Åsberg Depression Rating Scale; BDI, Beck Depression Inventory). **d)** All-night EEG spectral power density following placebo administration. Mean values are shown separately for NREM sleep (stages N2 and N3) and REM sleep. Blue-shaded regions indicate frequency bins (0.5 Hz resolution) that differed significantly between the two participant groups (t-test, p < 0.05). **e)** Slow-wave activity plotted in 30-second bins across 7.5 hours of sleep for a representative individual, shown across all three conditions (PLA: placebo; TRA: trazodone; GHB: gamma-hydroxybutyrate).

### Participants

All participants underwent a thorough assessment by an experienced study physician, covering general health and psychiatric history. The inclusion criteria for the MDD group were: a diagnosis of MDD according to the Diagnostic and Statistical Manual of Mental Disorders (DSM-5), stable antidepressant treatment (e.g., selective serotonin reuptake inhibitors [SSRIs] or serotonin–norepinephrine reuptake inhibitors [SNRIs]), age between 20 and 65 years, and no or low nicotine dependence, as indicated by a Fagerström Test for Nicotine Dependence total score of less than three. Exclusion criteria included the use of any sleep-promoting medication, such as benzodiazepines or z-drugs, on a regular basis or within three days before an experimental night; any axis-I DSM-5 psychiatric disorder other than MDD; neurological disorders or head injury; and any clinically relevant medical conditions. Detailed exclusion criteria can be found in the supplementary methods. The inclusion criteria for HC were: non-smoking, age between 20 and 65 years. Exclusion criteria were the same as in patients plus the presence of any axis-I DSM-5 psychiatric disorder.

### Intervention

Administration of study medications followed at two fixed times during each experimental night (Figure 1a-b). At 23:30, participants received two capsules containing either placebo (mannitol) or trazodone HCL (Trittico®) dosed at 1.5mg/kg (dose range: 72-156 mg), typical for hypnotic use compared to 300 mg/day for antidepressant use [27]. Trazodone was administered 30 minutes before bedtime, expected to reach peak concentrations after 1 hour [27]. At 3.30 h, participants were briefly awakened by an acoustic signal and minimal lighting to administer the liquid medication, either GHB (Xyrem®) or placebo (0.9% sodium chloride solution), mixed with 100 ml of orange juice to mask its salty taste. Administration took a median of 3 minutes (range: 1 to 10 min), including an optional toilet break. GHB was dosed according to body weight at 50 mg/kg (range 2400-5200 mg) and administered at 3:30 due to its short half-life and its slow-wave sleep enhancing effects seen in prior sleep studies [24]. This dosing regimen promotes sleep during a period when SWS is decreased due to reduced homeostatic sleep pressure, and sleep tends to be lighter, particularly in patients with hyperarousal, stress-related conditions, and early-morning awakening [28]. The effects of GHB on sleep physiology, brain connectivity, and stress biomarkers in healthy volunteers under this dosing regimen have been reported before [23] [24] [29] [30]. Both solid and liquid placebos were matched in appearance and taste to their respective medications. The liquid placebo was matched to GHB in appearance and taste and administered with 100 mL orange juice, whereas the solid placebo was provided in capsules identical in appearance to the active medication. All study medications were provided by the pharmacy of the University Hospital of Psychiatry Zurich, with balanced, block-wise randomization performed by an experienced pharmacist at the hospital’s pharmacy.

### Clinical assessment

Screening night: As depicted in Figure 1c, clinician-administered questionnaires included the Montgomery-Åsberg Depression Rating Scale (MADRS) and the Hamilton Depression Rating Scale (HAMD). Self-reported depressive symptoms were reported using Beck Depression Inventory (BDI). Substance use was assessed using the Alcohol Use Disorders Identification Test (AUDIT), Cannabis Use Disorders Identification Test (CUDIT), and Fagerström Test for Nicotine Dependence (FTND). Self-reported sleep quality was evaluated during the screening session using the Pittsburgh Sleep Quality Index (PSQI). Experimental nights: The Positive and Negative Affective Scale (PANAS) was used to assess momentary positive and negative affects both before (22:30) and after (09:30) the experimental night (24:00-07:30). Sleepiness was assessed using the Karolinska Sleepiness Scale (KSS) just before solid drug administration (23:30) and upon awakening (07:35). An evening questionnaire (23:30) was administered to record the intake of any caffeinated or alcoholic beverages, daytime naps, and medication use. Upon awakening, a morning questionnaire (MQ, 08:00) was used to evaluate self-reported sleep quality for each experimental night.

### Polysomnography

#### EEG setup

During the experimental nights, polysomnography (PSG) was recorded using an EEG amplifier (SIENNA Ultimate, EMS Handels GmbH, Austria) following the guidelines of the American Academy of Sleep Medicine (AASM) [31]. The setup included electroencephalography (EEG) with 20 scalp electrodes, two reference electrodes and a ground electrode placed according to the international 10-20 system, a bipolar electrooculogram, submental electromyogram, and a two-lead electrocardiogram (ECG). The analogue signals were conditioned by a high-pass filter (EEG: 0.5 Hz; EMG: 5 Hz; ECG: 1 Hz) and a low-pass filter (EEG: 70 Hz; EMG: 100 Hz; ECG: 70 Hz), and a notch-filter to remove line-noise (50 Hz), digitized and stored at the sampling frequency of 512 Hz.

#### Visual scoring

Sleep variables were visually scored based on 30-s epochs according to the criteria of the AASM using Rembrandt Analysis Manager (version 8, Embla Systems). For data scoring in 30-second epochs following AASM guidelines, the F3, C3, and O1 derivations were utilized, with F4, C4, and O2 used as alternatives in cases of severe artifacts. Two independent scorers evaluated the data, any discrepancies between the scorers were resolved by a third expert. The following sleep variables were calculated: (i) total sleep time (TST); (ii) sleep onset latency (SOL); (iii) wake after sleep onset (WASO); (iv) duration of sleep stages (i.e., non-REM [NREM] stage 1 [N1], NREM stage 2 [N2], NREM stage 3 [N3, SWS], NREM sleep [N2+N3], REM sleep); and (v) sleep efficiency ([SE] = [TST/time in bed] × 100). The duration of sleep stages is expressed as a percentage of TST.

#### EEG preprocessing

EEG data were preprocessed based on previously published procedures [25] [32]. In brief, signals were down-sampled to 128 Hz, bandpass filtered between 0.5–40 Hz and detrended. Muscle, ocular, ECG, and high-frequency EEG artifacts were identified using automated, threshold-based detectors applied to sleep-only data, and artifact-contaminated segments were removed [33]. Unreliable EEG snippets were interpolated. Electrical noise was attenuated using 50 Hz and 100 Hz notch filters together with frequency-domain suppression of device-specific harmonic peaks. A full description of all preprocessing steps, thresholds, and validation criteria is provided in the Supplementary Methods.

#### Quantitative EEG analysis

EEG data were segmented into 30-s epochs, and spectral power was computed from artifact-free data using Welch’s method. C3 spectral density was estimated in the 0.5–30 Hz range using 4-s Hanning windows with 50% overlap, and values were expressed in decibels [25]. Further methodological details are provided in the Supplementary Methods.

#### Spindle detection

Sleep spindles were automatically detected by using the *yasa.spindle_detect* function of the YASA algorithm [34], applied to artifact-free N2 and N3 sleep data from Fz and Cz EEG channels. Spindles of <100μV [35] and 0.5-3 s duration were detected using individualized detection thresholds (similar to [36]), with fixed frequency bands for slow (11–13 Hz) and fast (13–16 Hz) spindles, reflecting their frontal and central topographies. Spindle count, amplitude, frequency, and density per minute of NREM sleep, were calculated per individual, condition, and channel.

#### Slow-wave detection

Slow-waves were detected in Fz and Cz channels using the *yasa.sw_detect* function of the YASA algorithm during N2 and N3 sleep [34]. Candidate events were identified within the 0.5–1.25 Hz frequency range. For subsequent analyses, only strong slow oscillations were retained, defined as events whose peak-to-peak amplitude exceeded the participant-and channel-specific 75th percentile of the respective placebo condition [37].

#### Slow-oscillation – spindle co-occurrence and coupling

Temporal coupling between slow oscillations and spindles during NREM sleep was quantified using complementary event-based and phase-based metrics, based on the YASA algorithm [38].

##### Co-occurrence of discrete events

A spindle was considered co-occurring with a slow-oscillation (SS-SO) when its peak occurred within ±1.2 s of a slow-oscillation negative peak [39]. Conversely, a slow-oscillation was considered coupled when at least one spindle peak fell within ±1.2s of the slow-oscillation negative peak (SO-SS). For each participant, condition, and channel, the proportion of co-occurring spindles and slow oscillations was calculated relative to the total number of detected slow-oscillation events above the 75^th^ amplitude percentile [37].

##### Coupling phase

Preferred coupling phase was defined as the circular mean phase angle at which maximal sigma activity occurred relative to the slow oscillation cycle, averaged across co-occurring events for each participant, condition, and channel. The consistency of phase alignment across events was additionally quantified using the resultant vector length (RVL), with group differences in preferred phase assessed using circular statistics.

##### Coupling strength

Phase-amplitude coupling strength between instantaneous slow-oscillation phase and sigma-band amplitude was quantified using normalized direct phase-amplitude coupling (ndPAC) derived from YASA’s *yasa.sw_detect function* [40]. Mean phase-amplitude coupling strength was calculated per participant, condition, and channel across slow oscillations with a co-occurring discrete spindle event and non-zero ndPAC values included, with zero ndPAC indicating outlier data.

### Memory Tasks

#### Paired-Word Learning Task (PWLT)

The paired-word learning task was adapted from Gais and Born 2004 [41]. On each experimental night, participants were presented with one of three different word pair lists via Microsoft PowerPoint. Each word pair was shown for 800ms, preceded and followed by 200ms fixation crosses and a 2-second interstimulus interval. An immediate cued recall test followed in Matlab where the individual cueing words were presented sequentially on the screen and participants verbally recalled the associated word within a 5-second timeframe. Words that were not correctly recalled were re-presented for feedback for another 800ms. The following morning, participants were re-assessed with the same cued recall task, but without re-presenting missed words. For PWLT analyses, three participants were excluded due to issues with data acquisition (n=1 MDD, n=2 HC). Δaccuracy was calculated as the number of correctly recalled word pairs in the morning minus the number correctly recalled in the evening. Learned words were defined as word pairs recalled correctly only in the morning, whereas unlearned words were those recalled correctly only in the evening.

#### Finger Tapping Task (FTT)

The Sequential Finger Tapping Task [42] was administered via the computer software E-Prime, requiring participants to repeatedly execute one out of four five-digit sequences per experimental night using four keys (numbers 1-4) on the numeric keypad with their left (non-dominant) hand. The sequence remained visible on the monitor throughout the task to eliminate potential involvement of working memory. Following a practice trial (1-1-2-3-4), the actual sequence was encoded for 12 trials of 30s, each with a 30-second inter-trial interval. The following morning, after completing the short practice trial (1-1-2-3-4), 3 delayed recall trials of 30s followed. Participants were instructed to perform the task as quickly and accurately as possible. Three different outcome variables were calculated: Δaccuracy, Δcycles, and Δspeed, defined as the morning minus evening difference in the percentage of correctly tapped cycles, number of correctly tapped cycles, and number of tapped cycles per second, respectively.

#### Emotional Picture Memory Task (EPMT)

For the EPMT which was adapted from Rasch et al 2010 [43], pictures with positive, negative and neutral valence were chosen from the International Affective Picture System (IAPS). In total, 75 pictures were presented, with 25 images per positive, negative and neutral content category. Similar to Halonen et al 2023, pictures were presented to the participant for 2 seconds each [44]. After every 8 and 16 images, a 2-second pause was introduced, during which a white screen was displayed. After every 25 images, there was a 4-second pause. During the cued recall on the next morning, participants used a MATLAB program to distinguish, within 3s per picture, the 75 previously viewed images from 75 novel control images matched for visual content. Recognition sensitivity (d′) was calculated as the difference between the hit rate and false-alarm rate, with higher values indicating better discrimination between previously presented and novel pictures. Three different outcome variables were calculated: positive–neutral and negative–neutral scores, defined as the number of correctly recalled positive or negative pictures minus correctly recalled neutral pictures, respectively, and recognition sensitivity (d′), calculated as Z(hit rate) − Z(false-alarm rate).

### Statistical Analysis

Statistical analyses were conducted using R-Studio (version 2023.9.1.494, R-Studio Core Team). The significance level was set to p<0.05 (two-tailed). To assess model assumptions and goodness-of-fit, Normal Q–Q plots and Tukey-Anscombe (residuals vs. fitted) plots were examined. Variables with non-normal distributions were log-transformed or power-transformed as indicated. To investigate drug effects on outcomes, linear mixed-effects models (LMER) were used for continuous data, while generalized linear mixed-effects models of the Poisson family were used for count data (R-package *lme4*, Version 1.1.34). Condition (placebo vs. trazodone vs. GHB) and experimental night order were included as within-subject factors, group (HC vs. MDD) as between-subject factor, with subject ID as a random effect. After model fitting, analyses of variance (ANOVA) were performed to evaluate the overall significance of the fixed effects. If the condition significantly influenced an outcome variable, pairwise comparisons of estimated marginal means for each experimental condition were conducted using the *emmeans* package (Version 1.8.9). Post-hoc group-by-condition contrasts were performed only for outcomes showing a significant overall group × condition interaction. P-values from post-hoc comparisons were corrected for multiple comparisons using the FDR procedure.

Standardized effect sizes (ES) for post-hoc pairwise comparisons were calculated using the *eff_size* function of the *emmeans* package. ES values are derived from mean differences relative to the common standard deviation, with values greater than 0.8 or less than −0.8 considered to indicate large effects.

Circular (phase) data were analyzed separately using circular statistics implemented in the *circular* and *CircStats* packages. Within each group, non-uniformity of phase distributions was assessed using the Rayleigh test. Group differences and group-by-condition interactions in preferred phase were evaluated using the Watson–Williams test for independent samples. To assess within-subject condition effects, paired circular comparisons were conducted by computing angular differences between conditions and testing whether these were centered around zero using one-sample *V*-tests.

To examine associations between sleep parameters and memory variables, multiple linear regression analyses were conducted. All continuous predictors were z-standardized prior to analysis to obtain standardized regression coefficients (β). Models were adjusted for group effects.

## Results

### Participant characteristics

Of 31 patients with MDD initially assessed, 29 were randomized, 23 received at least one intervention, and 22 completed the study; of 25 HC assessed, 24 were randomized, 22 received at least one intervention, and 21 completed the protocol. One HC was excluded due to insufficient data resulting in 20 HC participants analyzed (see Supplementary Figure 1).

Baseline sociodemographic characteristics were comparable between groups (Table 1). MDD and HC did not differ in age, sex, body mass index (BMI), ethnicity, education, or alcohol and cannabis use (all p ≥ 0.071). At screening, the MDD group spent more time in bed (PSQI _Time in Bed,_ 8.8 ± 2.2 vs. 8.00 ± 0.6h, p = 0.016) and reported poorer subjective sleep quality (PSQI _Total score_) compared to the control group (7.1 ± 2.6 vs. 4.1 ± 0.95, p<0.001). Depression severity scores (MADRS, HAMD, BDI) were significantly higher in the MDD group (p _all_ < 0.001), see Figure 1c and Table 1.

**Table 1.** Sociodemographic and clinical characteristics at screening. Values are presented as mean ± standard deviation (SD) unless otherwise indicated. Education status (highest completed degree): I = primary school, II = lower secondary level, III = upper secondary level, IV = tertiary level. BMI = Body Mass Index; AUDIT = Alcohol Use Disorders Identification Test; CUDIT = Cannabis Use Disorders Identification Test; FTND = Fagerström Test for Nicotine Dependence; PSQI = Pittsburgh Sleep Quality Index; MADRS = Montgomery-Åsberg Depression Rating Scale; HAMD = Hamilton Depression Rating Scale; BDI = Beck Depression Inventory; SSRI = Selective Serotonin Reuptake Inhibitors; SNRI = Serotonin and Norepinephrine Reuptake Inhibitors; NDRI = Norepinephrine and Dopamine Reuptake Inhibitors; MDD = Major Depressive Disorder. Between-group comparisons were performed using either Welch’s t-tests or Mann-Whitney U tests.

|  | **MDD Mean** | **MDD SD** | **MDD Range** | **HC Mean** | **HC SD** | **HC Range** | **P<sub>t-test</sub>** |
|---|---|---|---|---|---|---|---|
| **Age, years** | 26.8 | 6.5 | 20–45 | 26.5 | 6.3 | 21–45 | 0.859 |
| **Sex, f/m** | 16 / 7 | - | - | 16 / 6 | - | - | 1 |
| **BMI, kg/m<sup>2</sup>** | 23.1 | 3.4 | 18.2–29.4 | 22.6 | 2.4 | 17.72–26.89 | 0.532 |
| **Ethnicity (self-reported), caucasian/asian/african/other** | 20/1/1/1 | - | - | 21/0/0/1 | - | - | 1 |
| **Education status, I/II/III/IV** | 0/1/15/7 | - | - | 0/0/9/13 | - | - | - |
| **AUDIT, score (alcohol)** | 3.6 | 3.3 | 0–15 | 3.1 | 2.1 | 0–8 | 0.514 |
| **CUDIT, score (cannabis)** | 1.7 | 2.3 | 0–7 | 0.7 | 1.3 | 0–3 | 0.071 |
| **FTND, score (nicotine)** | 1.7 | 0.5 | 1–2 | - | - | - | - |
| **PSQI time in bed, h** | 8.8 | 2.2 | 6–11 | 8.00 | 0.6 | 7.0–9.0 | 0.041 |
| **PSQI sleep latency, min** | 19.2 | 17.8 | 2–90 | 13.3 | 6.2 | 5–30 | 0.182 |
| **PSQI sleep efficiency, %** | 87.4 | 15.3 | 45.5–100 | 93.9 | 5.4 | 82.4–100 | 0.132 |
| **PSQI total, score** | 7.1 | 2.6 | 4–14 | 4.1 | 0.95 | 2–6 | <0.001 |
| **Medication, SSRI/SNRI/NDRI/vortioxetine/herbal** | 13/4/4/1/1 | - | - | 0/0/0/0/0 | - | - | - |
| **MDD episodes, n** | 4.1 | 4 | 1–20 | - | - | - | - |
| **MDD duration, years since first symptoms** | 8.1 | 5.7 | 1–24 | - | - | - | - |
| **MADRS, score** | 18.7 | 6.2 | 8–31 | 1.1 | 1.4 | 0–5 | <0.001 |
| **HAMD, score** | 17.4 | 5.4 | 8–29 | 1.1 | 1.6 | 0–5 | <0.001 |
| **BDI, score** | 19.2 | 8.8 | 2–37 | 1.6 | 2.9 | 0–13 | <0.001 |
| **Hospitalizations, n** | 1.3 | 1.8 | 0–6 | - | - | - | - |

### Electroencephalographic (EEG) sleep variables

#### Sleep stages

In the placebo condition, a higher proportion of NREM 1 (N1) sleep stage in MDD relative to HC was observed (p = 0.048, Cohen’s d = 0.62). There were no group differences in N2, N3 (also termed SWS), NREM (N2+N3), and rapid eye movement sleep (REM) stages, sleep onset latency (SOL), wake after sleep onset (WASO), total sleep time (TST), or sleep efficiency (SE) (all p ≥ 0.063).

In a linear mixed-effects model including fixed effects of condition, group, and night order, with a random intercept for participant, both drugs altered stage composition relative to placebo (see Table 2). GHB markedly increased N3 by +16.76% (p_FDR_ < 0.001, standardized effect size [ES] = 2.56) and NREM by +9.48% (p_FDR_ < 0.001, ES = 1.57), and decreased N1 (-2.56%, p_FDR_ < 0.001, ES= 1.24), REM (−7.40%, p_FDR_ < 0.001, ES = 1.29), and N2 (−7.26%, p_FDR_ < 0.001, ES= 1.16) sleep.

**Table 2.** Drug effects on nocturnal objective sleep variables. For each sleep variable, mean± standard deviation (SD) is provided separately for healthy controls (HC) and patients with major depressive disorder (MDD) across the three experimental conditions: placebo (PLA), trazodone (TRA), and gamma-hydroxybutyrate (GHB). The duration of sleep stages NREM 1 (N1), NREM 2 (N2), NREM 3 (N3), rapid eye movement (REM), and NREM sleep (N2 + N3) is expressed as a percentage of total sleep time (TST). Wake after sleep onset (WASO), sleep onset latency (SOL), and total sleep time (TST) are reported in minutes. Sleep efficiency (SE) is calculated as SE = (TST / total time in bed) × 100. Baseline group differences between HC and MDD during the placebo night were assessed using independent-samples t-tests or Wilcoxon rank-sum tests. PANOVA shows the condition effect from a linear mixed-effects model including condition and group as fixed factors and subject as a random intercept. Pairwise post hoc comparisons between conditions are reported based on estimated marginal means with false discovery rate correction.

|  | HC |  |  | MDD |  |  | P<sub>T-test/Wilcox</sub> | P<sub>Anova</sub> | P<sub>PLA-TRA</sub> | P<sub>PLA-GHB</sub> | P<sub>TRA-GHB</sub> |
|  | P | T | G | P | T | G | HC-MDD (PLA) | condition |  |  |  |
|---|---|---|---|---|---|---|---|---|---|---|---|
| **N1** | 5.13±3.09 | 5.44±3.53 | 3.88±2.53 | 7.06±4.04 | 8.62±6.71 | 4.43±3.83 | 0.048 | 0.005 | 0.520 | <0.0001 | <0.0001 |
| **N2** | 51.91±5.96 | 53.58±8.87 | 45.50±7.08 | 51.95±11.33 | 50.83±13.50 | 44.45±11.70 | 0.989 | <0.0001 | 0.862 | <0.0001 | <0.0001 |
| **N3** | 19.93±8.64 | 22.26±9.16 | 37.04±12.54 | 19.26±9.60 | 23.16±13.08 | 34.85±13.76 | 0.812 | <0.0001 | 0.052 | <0.0001 | <0.0001 |
| **NREM** | 71.84±5.26 | 75.83±5.15 | 82.54±7.02 | 71.21±7.54 | 73.99±9.24 | 79.30±8.99 | 0.754 | <0.0001 | 0.036 | <0.0001 | <0.0001 |
| **REM** | 23.03±4.66 | 18.73±4.18 | 13.58±5.81 | 21.73±7.03 | 17.39±8.06 | 16.27±8.96 | 0.484 | <0.0001 | 0.006 | <0.0001 | 0.012 |
| **SOL** | 19.52±16.75 | 24.10±22.15 | 21.73±14.01 | 23.07±18.66 | 24.68±16.52 | 16.62±10.23 | 0.357 | 0.606 | - | - | - |
| **WASO** | 19.90±17.26 | 22.78±23.45 | 21.20±20.04 | 38.07±43.82 | 15.88±13.48 | 18.81±14.90 | 0.063 | 0.980 | - | - | - |
| **TST** | 408±21 | 403±24 | 403±33 | 384±55 | 404±27 | 409±20 | 0.263 | 0.842 | - | - | - |
| **SE** | 91.31±4.87 | 89.72±5.97 | 90.38±5.89 | 86.30±12.07 | 90.96±5.64 | 92.13±3.88 | 0.230 | 0.701 | - | - | - |

Trazodone showed smaller effects, with a modest increase in NREM sleep of +3.00% (p_FDR_ = 0.036, ES = 0.50) and a reduction in REM sleep of −3.96% (p_FDR_ = 0.006, ES = 0.69). The increase in N3 under trazodone did not reach significance following FDR correction (+3.02%, p_FDR_ = 0.052, ES = 0.46).

Neither drug significantly altered sleep onset latency, wake after sleep onset, total sleep time, or sleep efficiency (all p ≥ 0.606), consistent with stage-specific effects rather than changes in overall sleep duration or continuity.

### Spectral variables

At baseline, few group differences in EEG spectral composition were observed. HC showed a larger NREM aperiodic exponent (p = 0.015, Cohen’s d = 0.78), increased NREM high-frequency delta power (d2; 2-4Hz; p = 0.002, Cohen’s d = 0.99), and increased REM high-frequency delta power (d2, 2-4 Hz; p = 0.006, Cohen’s d = 0.88). In contrast, patients with MDD, showed increased NREM (p = 0.004, Cohen’s d = 0.95) and REM (p = 0.026, Cohen’s d = 0.52) high-frequency beta power (b2; 20-30 Hz), see Figure 1d. No other placebo-night spectral differences were observed in the reported band-specific measures.

Drug administration induced marked frequency-specific changes in EEG spectral power. Across both study groups, GHB produced substantially larger spectral alterations than trazodone, with highly similar spectral profiles in patients with MDD and HC (s. Figure 2). GHB was characterized by increased low-frequency power together with reductions in higher-frequency activity during NREM sleep, whereas trazodone produced qualitatively similar but markedly smaller changes. Full spectral results are reported in Supplementary Table S1.

**Figure 2.**
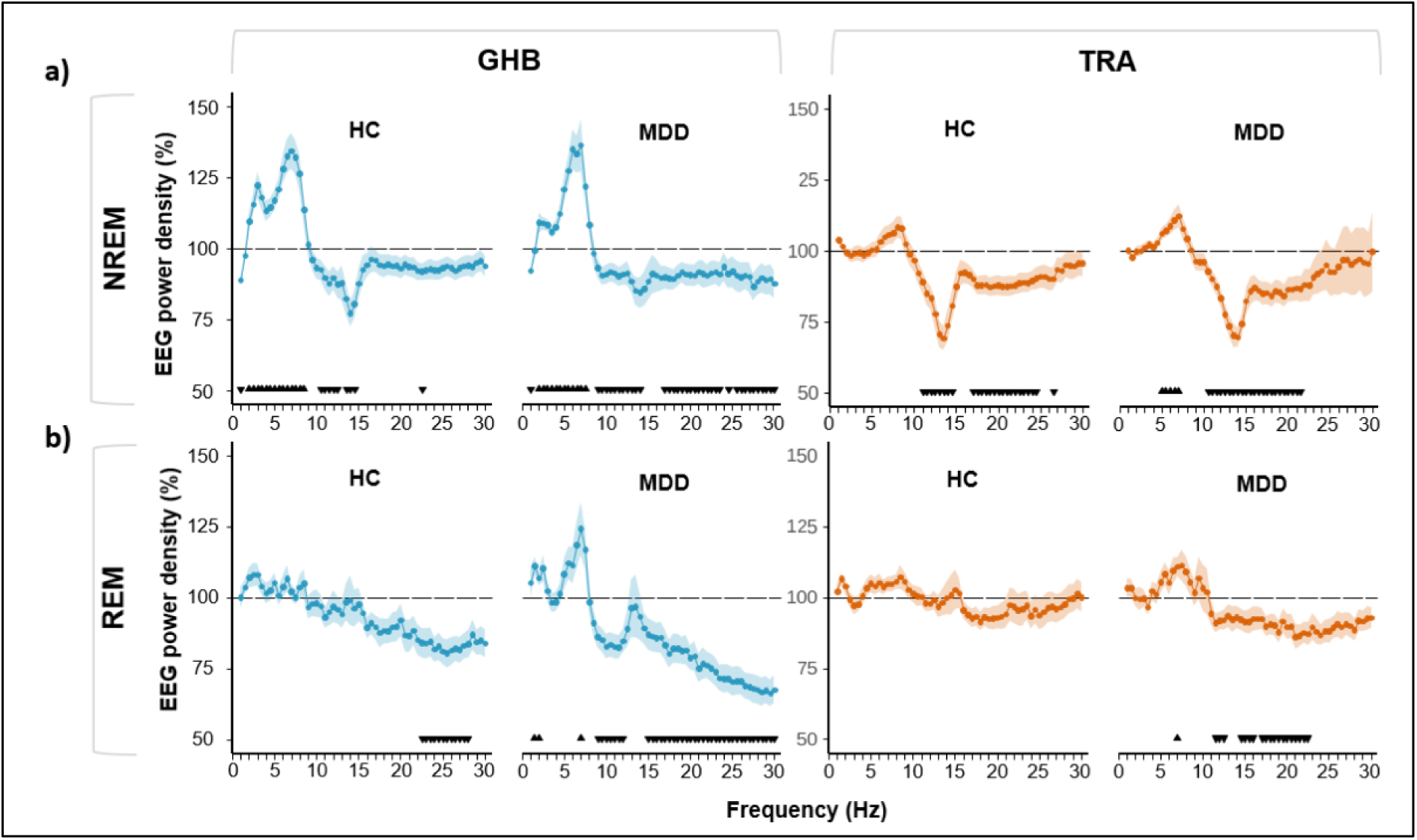
EEG power density. Whole-night spectral data from artifact-free epochs are shown for Healthy Controls (HC, left subpanels) and patients with Major Depressive Disorder (MDD, right subpanels) at the C3 electrode after administration of trazodone (TRA, 1.5 mg/kg) or solid placebo (PLA, 50mg/kg) at 23:30, and gamma-hydroxybutyrate (GHB) or liquid placebo (PLA) at 03:30. Power under each active condition is expressed relative to placebo, with placebo defined as 100% (dashed black line). **a)** The blue line illustrates placebo–GHB differences during NREM (upper left) and REM sleep (lower left). **b)** orange lines represent placebo–trazodone ratios during NREM (upper right) and REM sleep (lower right). Statistical comparisons were performed per frequency bin using t-tests. Significant differences are indicated by black triangles, whereby the direction of the triangle indicates an increase or decrease of power. Data are presented as mean ± SE. P-values were corrected for multiple comparisons using the Benjamini-Hochberg procedure.

### Spindle-slow oscillation temporal and phase coupling

#### Phase-based spindle–slow oscillation co-occurrence

Slow and fast spindles show distinct temporal profiles and topographical distribution[45]. The distributions between study groups (Figure 3a, upper panel) and between conditions (Figure 3a, lower panel) are shown for illustrative purposes.

**Figure 3.**
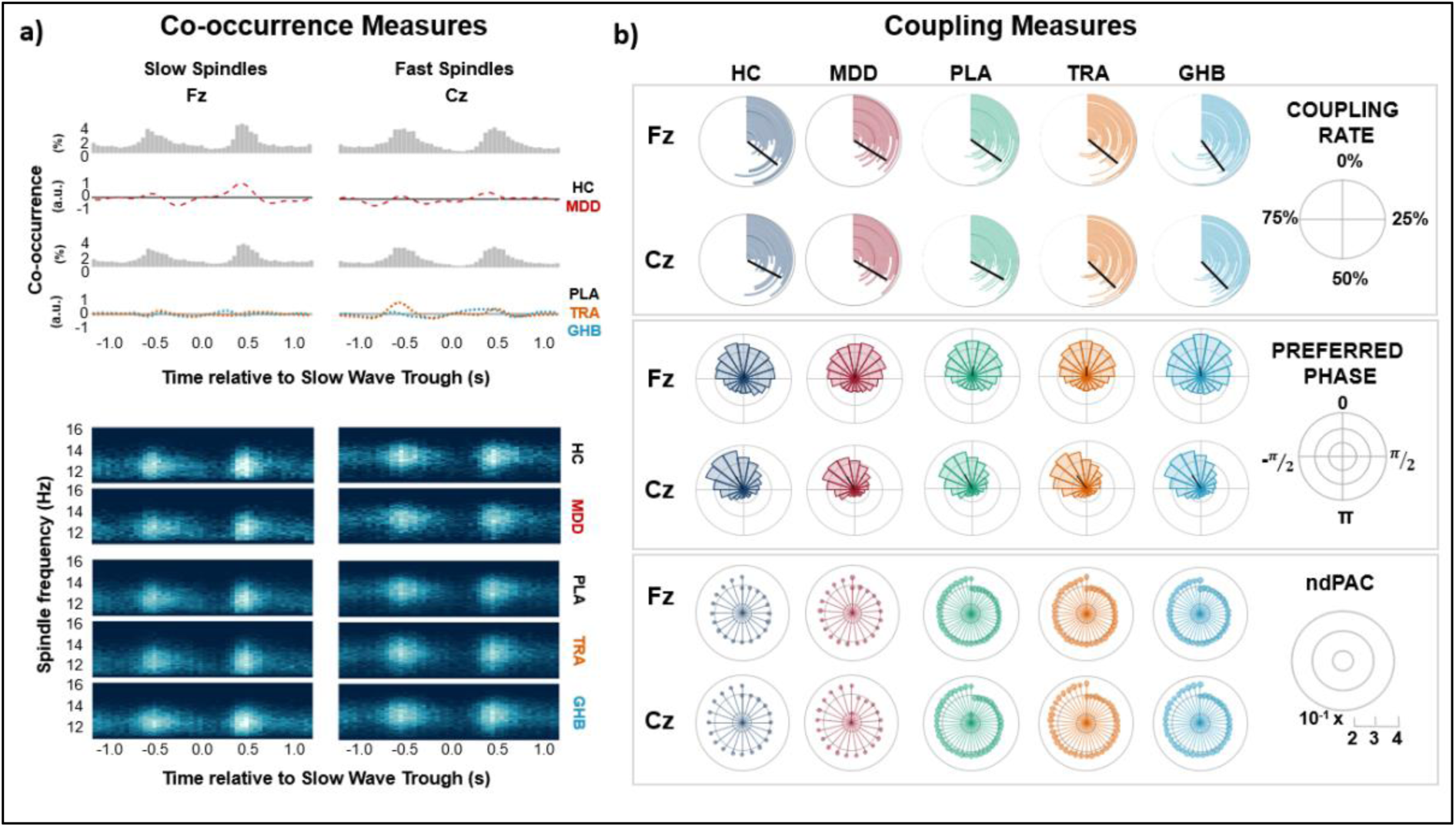
Visual representation of spindle measures and spindle – slow oscillation coupling centered around the negative slow oscillation peak during NREM sleep. **a)** Co-occurrence measures of spindles and slow oscillations (0.5-1.25 Hz) for slow (11-13 Hz, Fz, left column) and fast (13-16 Hz, Cz, right column) spindles at baseline between healthy control (HC, black line) and Major Depressive Disorder (MDD, red dashed line) groups (upper plot), as well as for condition effects (black line = placebo, orange = trazodone, blue = GHB) in pooled groups (lower plot) ± 1.2 seconds around the negative slow oscillation peak. The first row depicts a circular histogram of the percentage of spindles that occur together with a slow oscillation, with one value per individual. The middle row shows the difference in relative numbers between MDD (red dashed line) and HC (solid black line). The last row shows the heat map of spindle occurrence ± 1.2s around the negative slow oscillation peak (time = 0.0) for frequencies between 11-16 Hz per study group (upper panel) or condition (lower panel). **b)** Coupling measures for spindle – slow oscillation coupling. The two top rows show the co-occurrence rate of spindles with slow oscillations in Fz (upper row) and Cz (lower row) electrodes between groups during placebo (HC, dark blue; MDD, red) and for pooled groups (PLA, green; TRA, orange; GHB, blue). The two middle rows show distribution of preferred phase in 18×20° slices, with 0 being the positive peak of the slow oscillation and ±π being the negative slow oscillation peak. The lowest two rows show circular lollipop charts with one value per individual of phase-amplitude coupling strength (mean normalized direct phase-amplitude coupling; ndPAC) values, quantifying the strength of synchronization between SO phase and spindle-band amplitude within a ±1-s window around the SO negative peak.

#### Spindle–slow oscillation coupling

At Fz, HC showed higher slow-oscillation count (2663.1 ± 351.0 vs. 2222.0 ± 461.0; *p* = 0.001) and density (9.10 ± 1.09 vs. 8.33 ± 1.04; *p* = 0.003) than MDD, with no other baseline group differences.

Drug effects were pronounced (Figure 3b, top panel). Spindle count increased (PLA < TRA < GHB; all pairwise *p*FDR ≤ 0.010), whereas spindle density decreased (PLA > TRA > GHB; all pairwise *p*FDR ≤ 0.015). Slow-oscillation count increased (PLA < TRA < GHB; all *p*FDR < 0.001), and density was higher under GHB than PLA and TRA (both *p*FDR < 0.001). Both slow oscillations co-occurring with spindles and spindles co-occurring with slow oscillations increased in number from PLA to TRA to GHB (all pairwise *p*FDR < 0.001). Normalized direct phase-amplitude coupling (ndPAC), describing coupling strength between instantaneous slow-oscillation phase and sigma-band amplitude, was lower under GHB than TRA (*p*FDR = 0.033).

At Cz, HC likewise showed higher slow-oscillation count (2667.1 ± 355.1 vs. 2219.9 ± 465.7; *p*_FDR_ = 0.001) and density (9.12 ± 1.08 vs. 8.33 ± 1.03; *p*_FDR_ = 0.002) than MDD, with no other baseline group differences.

Spindle count increased (PLA < TRA < GHB; all pairwise *p*FDR ≤ 0.030), whereas spindle density decreased (PLA > TRA > GHB; all pairwise *p*FDR ≤ 0.024), while spindle amplitude was lower under both active conditions (both *p*FDR ≤ 0.022) and frequency was lower under GHB than PLA (*p*FDR = 0.0038). Slow-oscillation count and density increased across conditions (PLA < TRA < GHB; all *p*FDR ≤ 0.012), with higher amplitude under GHB than PLA and TRA (both *p*FDR ≤ 0.022). Both slow oscillations co-occurring with spindles and spindles co-occurring with slow oscillations increased in number from PLA to TRA to GHB (all pairwise *p*FDR < 0.001). Phase-amplitude coupling strength was higher under TRA than PLA (*p*FDR = 0.029).

Preferred spindle phase showed significant non-uniformity across all conditions and channels (all *p* < 0.001), clustering around 57–62° at Cz and 91–99° at Fz (Figure 3b, middle panel). No baseline HC–MDD differences were observed (Cz: *p*_FDR_ = 0.418; Fz: *p*_FDR_ = 0.996). Paired circular analyses indicated small condition-related phase differences at both channels (all *p*_FDR_ < 0.001): relative to PLA, preferred phase occurred earlier under TRA and GHB at Cz (5.9° and 2.5°) and Fz (6.8° and 3.9°), with smaller TRA–GHB differences (3.1° and 1.1°, respectively).

No baseline group differences were observed for coupling strength or consistency of phase alignment. Beyond the phase-amplitude coupling strength effects reported above, no other comparisons were significant, and consistency of phase alignment did not differ between conditions at either channel. More detailed information about coupling variables are shown in Supplementary Table S2.

#### Phase-based spindle–slow oscillation coupling

Preferred phase angles were consistently non-uniform across all conditions and channels (all *p* < 0.001), clustering around ∼57-62° at Cz and ∼91-99° at Fz, corresponding to a time point shortly after the slow-oscillation peak (Figure 3b, middle panel). Baseline group differences were evaluated under placebo using Watson–Williams tests for independent samples and revealed no differences between HC and patients with MDD at either Cz (*p*_FDR_ = 0.418) or Fz (*p*_FDR_ = 0.996).

Significant phase shifts were observed between all conditions at both channels (all *p*_FDR_ < 0.001), although the absolute changes in preferred phase were small. At Cz, preferred phase occurred earlier under trazodone and GHB than under placebo (PLA-TRA: 5.9°; PLA-GHB: 2.5°), whereas GHB occurred slightly later than trazodone (3.1°). At Fz, preferred phase also shifted earlier under trazodone and GHB relative to placebo (PLA-TRA: 6.8°; PLA-GHB: 3.9°), with a small difference between GHB and trazodone (1.1°). These findings indicate consistent but subtle within-participant rotations of preferred spindle phase across pharmacological conditions in the absence of baseline group differences.

No baseline group differences were observed for coupling strength or consistency of phase alignment. Drug-related differences in coupling strength were observed between placebo and trazodone at Cz (*p*_FDR_ = 0.029) and between trazodone and GHB at Fz (*p*_FDR_ = 0.033), whereas all other coupling strength comparisons were non-significant (all *p*_FDR_ ≥ 0.175). Consistency of phase alignment did not differ between conditions at either channel (all *p*_FDR_ ≥ 0.140).

#### Declarative, procedural and emotional memory

##### Paired Word Learning Task (PWLT)

Memory performance was quantified by overnight change including number of learned words, number of forgotten words, and the difference in accuracy between morning and evening recall. Under placebo, HC showed stronger overnight improvement in accuracy compared with MDD (10.47 ± 4.44 vs. 7.43 ± 4.69, p = 0.042, Cohen’s d = 0.65), while the number of learned (11.74 ± 4.21 vs. 9.19 ± 3.91) and forgotten (1.26 ± 1.1 vs. 1.76 ± 1.67) words did not differ by group (all p ≥ 0.056), see Figure 4a. In a linear mixed-effects model including fixed effects of condition, group, and night order, with a random intercept for participant, no significant effects of condition on PWLT performance were observed after FDR correction. Participants showed a tendency toward increased forgetting under GHB compared with placebo (Δ = −0.323, p_FDR_ = 0.094, ES =-0.32), whereas forgetting under trazodone did not differ from placebo (Δ = +0.084, p_FDR_ = 0.661, ES = 0.08). Immediate learning and overall change in accuracy were not affected by condition (all p_FDR_ ≥ 0.59). No significant group-condition interactions were observed (all pFDR ≥.757).

**Figure 4.**
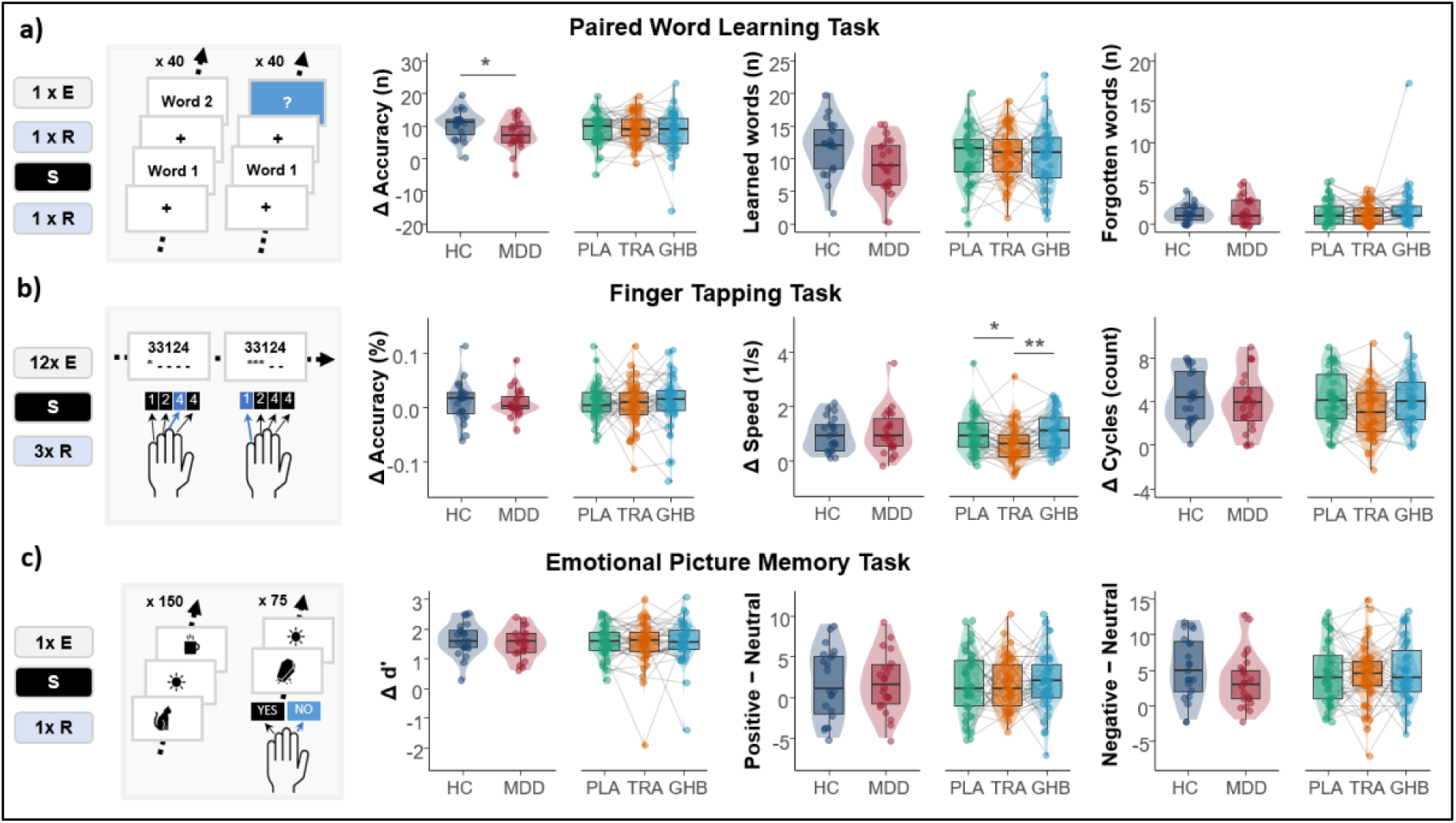
**Memory consolidation tasks design and results**. **a)** Paired Word Learning Task (PWLT). In the evening, participants learned a list of 40 word pairs (encoding [E]), followed by an immediate recall test (R), before a 7.5-h sleep opportunity (S). The following morning, 115 minutes after awakening, cued recall was performed. ΔAccuracy depicts the difference of correctly recalled words in the morning minus correctly recalled words in the evening. Left subpanels show baseline differences between healthy controls (dark blue) and the MDD group (red); right subpanels show condition effects across all participants for the three treatment nights (placebo = green, trazodone = orange, GHB = blue). **b)** Finger Tapping Task (FTT). Participants encoded a five-digit sequence with the non-dominant hand across 12 evening trials of 30 seconds each, followed by three recall trials the next morning 140 min after awakening. ΔAccuracy describes the difference of correctly tapped sequences in the morning minus correctly tapped sequences in the evening. **c)** Emotional Picture Memory Task (EPMT). In the evening, participants viewed 75 pictures (25 negative, 25 neutral, 25 positive). Morning recall was conducted 125 min after awakening and consisted of recognition memory for the 75 previously seen images and 75 novel foil images matched for content.

##### Finger Tapping Task (FTT)

Performance was assessed as overnight changes in accuracy, number of completed cycles, and speed.^42^ Under placebo, groups did not differ in overnight changes in accuracy, number of completed cycles, or speed (all p ≥ 0.426), see Figure 4b. In a linear mixed-effects model including fixed effects of condition, group, and night order, with a random intercept for participant, overnight motor sequence improvement was similar across conditions for accuracy and number of completed cycles (all p_FDR_ ≥ 0.078). However, trazodone was associated with reduced overnight speed gains compared with both placebo (Δ = 0.320, p_FDR_ = 0.012, ES = 0.62) and GHB (Δ = −0.370, p_FDR_ = 0.0076, ES =-0.72). No difference was observed between placebo and GHB (p_FDR_ = 0.67). No significant group-condition interactions were observed (all *p*_FDR_ ≥.152).

##### Emotional Picture Memory Task (EPMT)

Memory performance was quantified by sensitivity (d′) and emotional difference scores (number of recalled positive minus neutral pictures, and number of recalled negative minus neutral pictures).^4344^ Under placebo, MDD and HC did not differ in any of the scores (all p ≥ 0.191), see Figure 4c. In a linear mixed-effects model including fixed effects of condition, group, and night order, with a random intercept for participant, neither GHB nor trazodone influenced emotional difference scores or d’ (all p ≥ 0.268). A significant group-condition interaction was observed for TRA–GHB for negative relative to neutral emotional memory performance (*p*_FDR_ = 0.031, ES = −1.19).

#### Associations between sleep and memory variables

As an exploratory analysis, we tested associations between macrostructural, spectral and event-based polysomnographic measures and overnight memory performance, to explore which sleep physiological patterns best predict memory consolidation (see Figure 5). Among macrostructural measures, longer sleep onset latency and a higher proportion of N2+N3 sleep were associated with more forgotten words on the PWLT (β =-0.20, p = 0.038; β = 0.22, p = 0.039). Emotional memory bias was related to sleep-stage composition, with higher N1 and N2 percentages associated with larger EPMT negative-neutral differences (β = 0.24, p = 0.013; β = 0.19, p = 0.048), and higher N3 percentage associated with smaller differences (β =-0.28, p = 0.010). Greater total sleep time was associated with lower EPMT positive-neutral scores (β =-0.19, p = 0.041).

**Figure 5.**
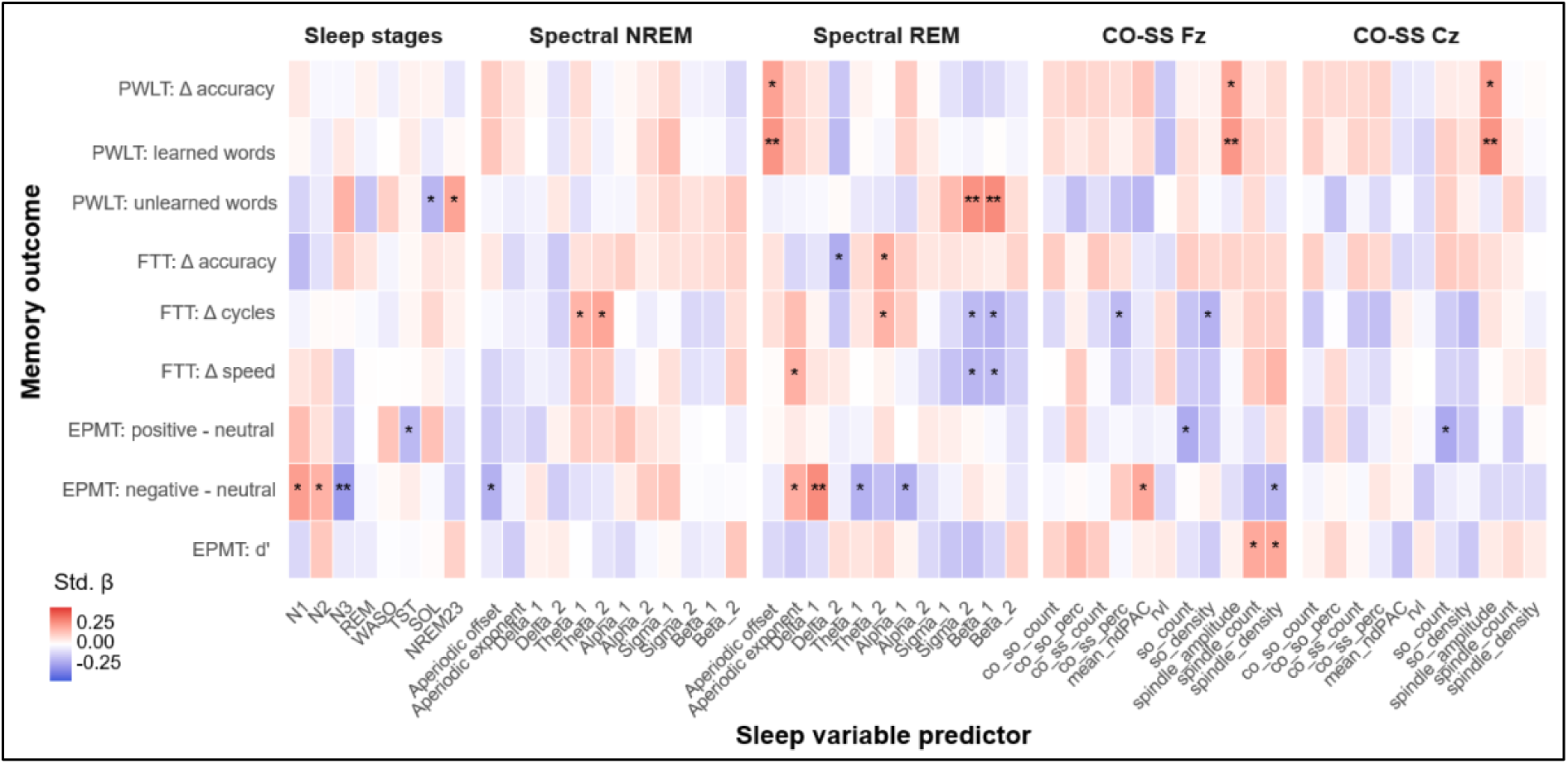
Associations between sleep-derived predictors and overnight memory variables. The heat map shows standardized regression coefficients for associations of macrostructural, spectral and spindle–slow-oscillation measures with next-morning memory variables across all experimental conditions. Values printed in each cell represent standardized beta coefficients. * = p<0.05; ** p<0.01. co_so_count, count of slow oscillations with co-occurring spindle; co_so_perc, percentage of slow oscillations with co-occurring spindle; co_ss_count, count of spindles with co-occurring slow oscillation; co_ss_perc, percentage of spindles with co-occurring slow oscillation; Cz, Cz EEG electrode; EPMT, Emotional Picture Memory Task; EPMT positive-neutral, number of correctly recalled pictures with positive content minus neutral content; EPMT negative-neutral, number of correctly recalled pictures with negative content minus neutral content; EPMT d’, Recognition sensitivity, calculated as hit rate - false-alarm rate; FTT, Finger Tapping Task; FTT Δaccuracy, percentage of correctly tapped cycles in the morning minus evening; FTT Δcycles, number of correctly tapped cycles in the morning minus evening; FTT Δspeed, number of tapped cycles per second in the morning minus evening; Fz, Fz EEG electrode; mean_ndPAC, mean Normalized direct phase-amplitude coupling; N1-N3, Non-REM stage 1-3; NREM23, Non-rem sleep stages 2 + 3; PWLT, Paired-Word Learning Task; PWLT Δaccuracy, number of correctly recalled word-pairs in the morning minus number of correctly recalled words in the evening; PWLT learned words, word pairs correctly recalled only in the morning; PWLT unlearned words, word pairs correctly recalled only in the evening, REM, REM sleep; SOL, sleep onset latency; TST, total sleep time; WASO, wake after sleep onset. Spectral power bands of the EEG: delta 1 (0.5-1 Hz), delta 2 (2-4 Hz), theta 1 (4-6 Hz), theta 2 (6-8 Hz), alpha 1 (8-10 Hz), alpha 2 (10-12 Hz), sigma 1 (11-13 Hz), sigma 2 (13-16 Hz), beta 1 (15-20 Hz), beta 2 (20-30 Hz).

Spectral associations emerged in both NREM and REM sleep. In NREM sleep, higher theta-1 (4-6 Hz) and theta-2 (6-8 Hz) power were associated with greater overnight improvement in FTT cycles (β = 0.18, p = 0.041; β = 0.22, p = 0.017), while higher aperiodic offset was associated with lower EPMT negative-neutral scores (β =-0.21, p = 0.030). In REM, higher aperiodic offset was associated with better PWLT performance (delta accuracy: β = 0.23, p = 0.022; learned words: β = 0.26, p = 0.009). Higher beta-1 (15-20 Hz) and sigma-2 (13-16 Hz) power were associated with more PWLT forgetting (β = 0.28, p = 0.003; β = 0.27, p = 0.004) and poorer FTT performance (cycles: β =-0.20, p = 0.029; speed: β =-0.19, p = 0.041; cycles: β =-0.19, p = 0.039; speed: β =-0.20, p = 0.027). Emotional memory was also linked to REM spectral features, with alpha-1 (8-10 Hz) and theta-1 (4-6 Hz) power associated with lower EPMT negative-neutral scores (β =-0.21, p = 0.021; β =-0.21, p = 0.025), and aperiodic exponent and delta-1 (0.5-1Hz) power associated with higher negative-neutral scores (β = 0.20, p = 0.040; β = 0.28, p = 0.002).

Event-based associations were more selective. Lower frontal spindle co-occurrence percentage and slow-oscillation density were associated with reduced FTT cycle improvement (β =-0.19, p = 0.048; β =-0.21, p = 0.037). Frontal coupling strength was positively associated with EPMT negative-neutral scores (β = 0.21, p = 0.026), whereas higher slow-oscillation count was associated with lower EPMT positive-neutral scores (β =-0.23, p = 0.037). Spindle amplitude at both Fz and Cz was positively associated with PWLT performance (delta accuracy: β = 0.22, p = 0.017; learned words: β = 0.25, p = 0.007; delta accuracy: β = 0.23, p = 0.013; learned words: β = 0.26, p = 0.005). Additionally, higher frontal spindle count and density were associated with higher EPMT d′ (β = 0.21, p = 0.027; β = 0.21, p = 0.027), while higher frontal spindle density and central slow-oscillation count were associated with lower emotional memory bias (β =-0.19, p = 0.044; β =-0.23, p = 0.034).

## Discussion

In this randomized, double-blind, placebo-controlled crossover trial, nocturnal administration of GHB increased SWS in both patients with MDD and HC, whereas trazodone produced smaller changes in sleep-stage composition. Contrary to our hypothesis, however, we observed no improvements in declarative, procedural, or emotional memory consolidation under either pharmacological condition.

These results challenge the assumption that increasing SWS is sufficient to enhance memory consolidation [9]. Although NREM sleep is closely linked to hippocampus-dependent memory processes [46], and synaptic plasticity [4], pharmacological sleep enhancement may increase the quantity of SWS without fully reproducing the physiological organization of sleep that supports systems memory consolidation [11]. Indeed, GHB strongly increased slow-oscillation density while slightly reducing spindle count, density and amplitude, mirroring a dissociation previously described for tiagabine [10], which enhanced slow-wave activity but suppressed slow-oscillation-locked spindles without benefiting memory consolidation. Notably, although both drugs promoted sleep more strongly in patients than in controls, this did not translate into a normalization of memory: even declarative consolidation, the domain in which patients differed from controls under placebo, showed no improvement under GHB, and no group-by-condition interaction emerged for any memory variables compared to placebo. At baseline, patients with MDD showed reduced slow-oscillation density, a trend towards lower spindle count, reduced delta power and spectral slope, and increased beta power and light sleep, consistent with previous work [47] [48]. However, measures of slow oscillation–spindle coupling differed only modestly between groups. The dissociation between altered sleep composition and oscillatory expression, on the one hand, and relatively intact coupling measures, on the other, further supports the idea that simple enhancement of SWS is unlikely to show large effects on the restoration of memory function [12].

In line with this interpretation, exploratory analyses across experimental conditions revealed only weak and inconsistent associations between global sleep-stage measures and next-day memory performance. In contrast, several microstructural sleep physiology measures showed more consistent relationships. Higher spindle amplitude was consistently associated with better declarative memory, while spindle density and slow oscillation density were linked to aspects of emotional and procedural memory. Spectral features also showed distinct associations with learning and forgetting, potentially reflecting differences in cortical excitation–inhibition dynamics relevant to sleep-dependent memory processing. By contrast, slow oscillation–spindle coupling exhibited only limited associations with behavioral variables. Together, these findings suggest that memory consolidation depends less on the overall quantity of SWS than on the integrity and expression of specific sleep oscillations embedded within it [5]. Rather than globally increasing SWS, interventions that selectively modulate functionally relevant oscillatory features, including spindle expression and the temporal organization of sleep rhythms, may represent a more promising strategy.

The clinical implications are particularly relevant for MDD. Sleep disturbances are core features of the disorder, and targeting sleep has been proposed as a strategy to improve cognition [49]. Our findings do not support the idea that promoting SWS is sufficient to enhance memory consolidation. Other downstream mechanisms, including altered synaptic plasticity, motivational factors or affective biases, may play a more prominent role.

Several limitations should be considered. The sample size was sufficient to detect moderate-to-large effects on sleep physiology but may have limited the detection of smaller effects on memory variables. Consequently, the study was also not powered to investigate individual differences in treatment response. Differences in pharmacokinetics and dosing schedules between GHB and trazodone limit direct comparisons between the compounds. However, the timing of drug administration was selected to maximize SWS enhancement for each compound, thereby reflecting their optimal clinical use rather than enforcing matched pharmacokinetic profiles. In addition, the memory battery focused on established paradigms of declarative, procedural, and emotional memory and did not incorporate targeted memory reactivation or other cueing paradigms during sleep, which may facilitate memory consolidation under experimental conditions but currently have limited applicability in routine clinical practice [50]. Finally, the study examined the effects of a single night of pharmacological intervention. Although repeated administration may yield different variables, the single-dose design provides a clear mechanistic test of whether pharmacological enhancement of SWS is sufficient to influence overnight memory consolidation. At the same time, the study combines robust pharmacological manipulation across two compounds with detailed electrophysiological characterization and multimodal assessment of memory consolidation in both patients with MDD and HC.

In summary, pharmacological modulation of SWS with GHB and trazodone did not improve memory consolidation in patients with MDD and HC. The dissociation between strong SWS changes with GHB and lack of effects on memory performance suggests that targeting SWS alone is unlikely to yield cognitive benefits. Future work should focus on interventions that more directly modulate sleep microstructure or combine sleep-based approaches with strategies targeting plasticity and memory.

## Additional Information

### Author contributions

LKS: Formal analysis; Investigation; Data Curation; Writing - Original Draft; Visualization; Project administration. BBQ: Conceptualization; Methodology; Writing - Review & Editing; Supervision. BS: Methodology; Formal analysis. JM: Formal analysis. JDF: Investigation; Data Curation. BR: Review & Editing; Supervision. DAD: Conceptualization; Investigation; Writing - Review & Editing. OGB: Conceptualization; Methodology; Writing - Review & Editing; Supervision; Funding acquisition. ES: Conceptualization; Resources; Writing - Review & Editing; Supervision; Funding acquisition. HPL: Conceptualization; Resources; Methodology; Writing - Review & Editing; Supervision. FB: Conceptualization; Methodology; Formal analysis; Investigation; Data Curation; Writing - Original Draft; Visualization; Project administration.

## Data Availability

All data produced in the present study are available upon reasonable request to the authors.

## Acknowledgements

We thank Lhanzom Lhasam, Jacqueline Kientsch, Marina Janki, Viveka Boller, Nicole Friedli, Yara Kreis, Anna Habereder, Rahel Gelin, Thomas Monasterios Gallardo, Zoe Marti, and Corinne Eicher for the support with data collection and analysis, Helena Jenzer and Monika Visentini for the support with study planning.

## Supplementary Figures

**Supplementary Figure S1.**
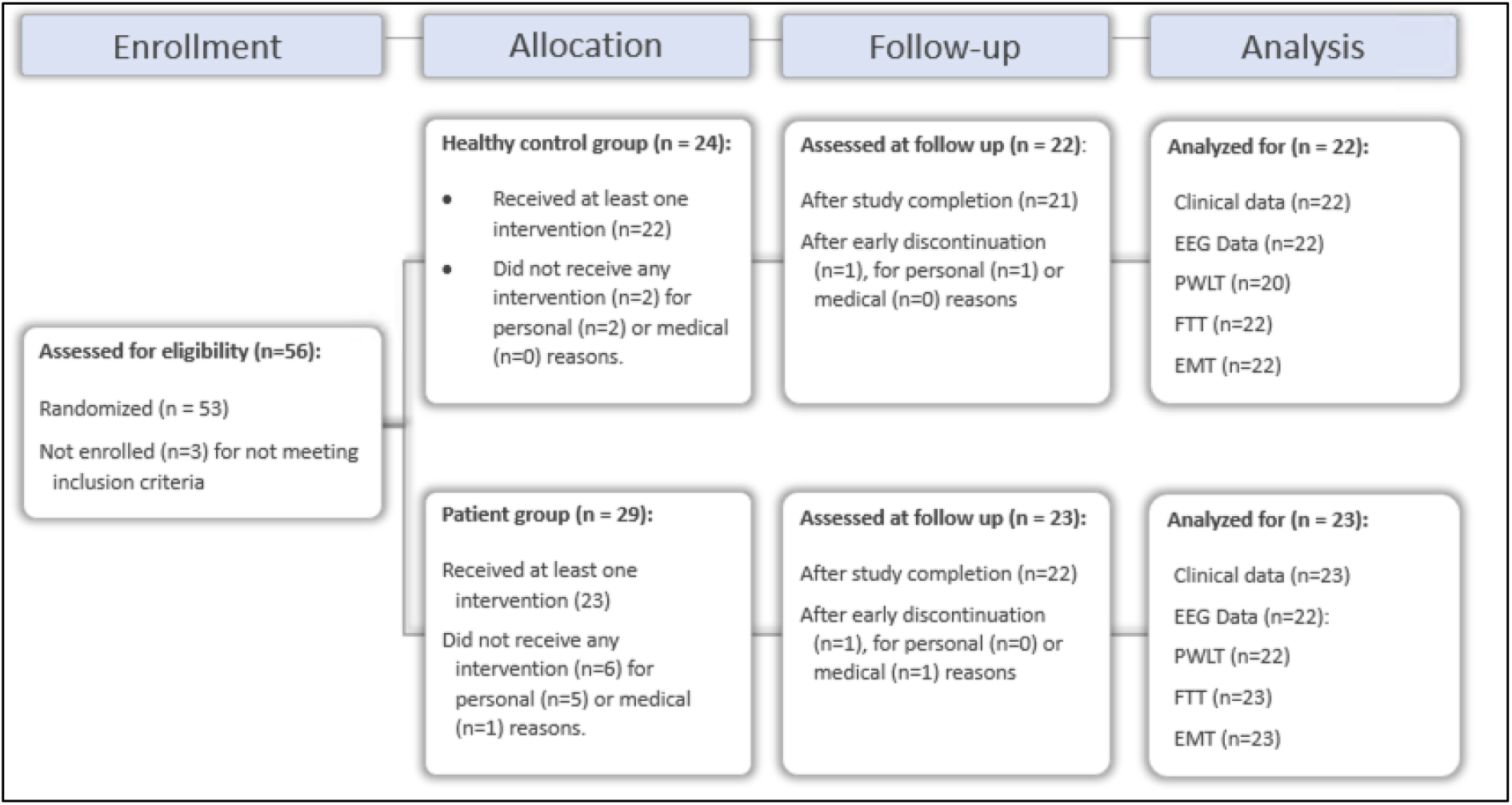
**Study protocol CONSORT diagram**. A total of 56 participants were assessed for eligibility, of whom 53 were enrolled in the study. Twenty-four participants were allocated to the healthy control group; 22 received at least one intervention and were included in the data analysis, and 21 completed the study. Two controls discontinued participation after enrollment but prior to the first experimental night due to personal reasons (study protocol being too time-consuming, *n* = 2), and one withdrew during the study because of adverse events. Twenty-nine participants were allocated to the patient group; 23 received at least one intervention. Six patients discontinued after enrollment but before the first experimental night for personal (feeling anxious/unwell in the sleep laboratory environment, *n* = 2; study protocol being too time-consuming, *n* = 2) or medical reasons (newly emerging medical conditions, *n* = 2). No patient withdrew due to adverse events. One patient reported amphetamine use and subsequent clinical worsening a few days after the first experimental night (trazodone) and withdrew from the study.

## Supplementary Tables

**Supplementary Table S1.** Spectral power and aperiodic component statistics during REM and NREM sleep. The table summarizes group differences and condition effects on relative EEG spectral power (0.5-30 Hz) as well as aperiodic parameters (*aperiodic exponent* and *aperiodic offset*) during REM and NREM sleep. Results are shown separately for healthy controls (HC) and patients with major depressive disorder (MDD). Between-group differences within each condition (PLA, TRA, GHB) were assessed using independent-samples *t*-tests or Wilcoxon rank-sum tests. Effects of condition are reported in the column “P_Anova_”. Post-hoc pairwise comparisons between conditions were performed using estimated marginal means. Statistical significance was defined as *p* < 0.05. Spectral power bands of the EEG: delta 1 (0.5-1 Hz), delta 2 (2-4 Hz), theta 1 (4-6 Hz), theta 2 (6-8 Hz), alpha 1 (8-10 Hz), alpha 2 (10-12 Hz), sigma 1 (11-13 Hz), sigma 2 (13-16 Hz), beta 1 (15-20 Hz), beta 2 (20-30 Hz).

|  |  | **HC** |  |  | **MDD** |  |  | P<sub>T-test/Wilcox</sub> | P<sub>Anova</sub> | P<sub>PLA-TRA</sub> | P<sub>PLA-GHB</sub> | P<sub>TRA-GHB</sub> |
|  |  | **P** | **T** | **G** | **P** | **T** | **G** | **HC-MDD (PLA)** | **condition** |  |  |  |
|---|---|---|---|---|---|---|---|---|---|---|---|---|
| **REM** | **d1** | 0.33 ± 0.05 | 0.33 ± 0.04 | 0.31 ± 0.04 | 0.34 ± 0.08 | 0.35 ± 0.05 | 0.34 ± 0.08 | 0.655 | 0.711 | - | - | - |
|  | **d2** | 0.27 ± 0.03 | 0.27 ± 0.03 | 0.29 ± 0.04 | 0.24 ± 0.04 | 0.24 ± 0.05 | 0.25 ± 0.04 | 0.006 | 0.009 | 0.608 | 0.023 | 0.011 |
|  | **t1** | 0.15 ± 0.02 | 0.15 ± 0.03 | 0.15 ± 0.02 | 0.14 ± 0.02 | 0.14 ± 0.02 | 0.14 ± 0.02 | 0.141 | 0.613 | - | - | - |
|  | **t2** | 0.09 ± 0.02 | 0.09 ± 0.02 | 0.09 ± 0.01 | 0.09 ± 0.02 | 0.09 ± 0.02 | 0.10 ± 0.04 | 0.294 | 0.733 | - | - | - |
|  | **a1** | 0.06 ± 0.02 | 0.06 ± 0.02 | 0.06 ± 0.02 | 0.07 ± 0.03 | 0.06 ± 0.02 | 0.06 ± 0.03 | 0.422 | 0.700 | - | - | - |
|  | **a2** | 0.03 ± 0.01 | 0.03 ± 0.01 | 0.03 ± 0.01 | 0.03 ± 0.01 | 0.03 ± 0.01 | 0.03 ± 0.01 | 0.517 | 0.173 | - | - | - |
|  | **s1** | 0.02 ± 0.01 | 0.02 ± 0.01 | 0.02 ± 0.01 | 0.03 ± 0.01 | 0.02 ± 0.01 | 0.02 ± 0.01 | 0.285 | 0.167 | - | - | - |
|  | **s2** | 0.02 ± 0.00 | 0.02 ± 0.01 | 0.02 ± 0.01 | 0.03 ± 0.01 | 0.02 ± 0.01 | 0.02 ± 0.01 | 0.194 | 0.544 | - | - | - |
|  | **b1** | 0.04 ± 0.01 | 0.04 ± 0.01 | 0.04 ± 0.01 | 0.05 ± 0.02 | 0.04 ± 0.01 | 0.04 ± 0.02 | 0.072 | 0.201 | - | - | - |
|  | **b2** | 0.03 ± 0.01 | 0.02 ± 0.01 | 0.02 ± 0.01 | 0.04 ± 0.02 | 0.03 ± 0.01 | 0.02 ± 0.01 | 0.026 | 0.046 | 0.002 | <0.001 | 0.011 |
|  | **Aperiodic exponent** | 1.64 ± 0.13 | 1.67 ± 0.11 | 1.72 ± 0.14 | 1.57 ± 0.17 | 1.61 ± 0.13 | 1.74 ± 0.19 | 0.137 | 0.001 | 0.026 | <0.001 | <0.001 |
|  | **Aperiodic offset** | 1.26 ± 0.18 | 1.29 ± 0.19 | 1.40 ± 0.19 | 1.22 ± 0.20 | 1.26 ± 0.19 | 1.45 ± 0.17 | 0.496 | <0.001 | 0.060 | <0.001 | <0.001 |
| **NREM** | **d1** | 0.56 ± 0.05 | 0.58 ± 0.05 | 0.54 ± 0.04 | 0.57 ± 0.07 | 0.59 ± 0.07 | 0.58 ± 0.07 | 0.518 | 0.002 | 0.004 | 0.974 | 0.004 |
|  | **d2** | 0.21 ± 0.03 | 0.21 ± 0.02 | 0.23 ± 0.03 | 0.18 ± 0.02 | 0.18 ± 0.03 | 0.19 ± 0.03 | 0.002 | <0.001 | 0.422 | <0.001 | <0.001 |
|  | **t1** | 0.08 ± 0.01 | 0.08 ± 0.01 | 0.08 ± 0.01 | 0.07 ± 0.01 | 0.08 ± 0.02 | 0.08 ± 0.02 | 0.486 | 0.111 | - | - | - |
|  | **t2** | 0.05 ± 0.01 | 0.05 ± 0.02 | 0.05 ± 0.02 | 0.05 ± 0.02 | 0.05 ± 0.02 | 0.06 ± 0.02 | 0.250 | <0.001 | 0.035 | <0.001 | 0.005 |
|  | **a1** | 0.03 ± 0.01 | 0.03 ± 0.01 | 0.03 ± 0.01 | 0.04 ± 0.02 | 0.03 ± 0.02 | 0.03 ± 0.01 | 0.330 | 0.041 | 0.624 | <0.001 | <0.001 |
|  | **a2** | 0.02 ± 0.01 | 0.02 ± 0.01 | 0.02 ± 0.01 | 0.02 ± 0.01 | 0.02 ± 0.01 | 0.02 ± 0.01 | 0.250 | <0.001 | <0.001 | <0.001 | 0.004 |
|  | **s1** | 0.02 ± 0.01 | 0.02 ± 0.01 | 0.02 ± 0.01 | 0.03 ± 0.01 | 0.02 ± 0.01 | 0.02 ± 0.01 | 0.452 | <0.001 | <0.001 | <0.001 | 0.951 |
|  | **s2** | 0.03 ± 0.01 | 0.02 ± 0.01 | 0.02 ± 0.01 | 0.03 ± 0.01 | 0.02 ± 0.01 | 0.02 ± 0.01 | 0.715 | <0.001 | <0.001 | <0.001 | 0.908 |
|  | **b1** | 0.04 ± 0.01 | 0.03 ± 0.01 | 0.03 ± 0.01 | 0.04 ± 0.02 | 0.03 ± 0.01 | 0.03 ± 0.01 | 0.996 | <0.001 | <0.001 | <0.001 | 0.754 |
|  | **b2** | 0.01 ± 0.00 | 0.01 ± 0.00 | 0.00 ± 0.00 | 0.01 ± 0.00 | 0.01 ± 0.00 | 0.01 ± 0.00 | 0.004 | <0.001 | <0.001 | <0.001 | 0.043 |
|  | **Aperiodic exponent** | 2.35 ± 0.11 | 2.41 ± 0.10 | 2.48 ± 0.14 | 2.23 ± 0.19 | 2.32 ± 0.18 | 2.39 ± 0.16 | 0.015 | <0.001 | <0.001 | <0.001 | <0.001 |
|  | **Aperiodic offset** | 2.20 ± 0.21 | 2.29 ± 0.21 | 2.40 ± 0.25 | 2.12 ± 0.24 | 2.21 ± 0.24 | 2.30 ± 0.24 | 0.222 | <0.001 | <0.001 | <0.001 | <0.001 |

**Supplementary Table S2.**
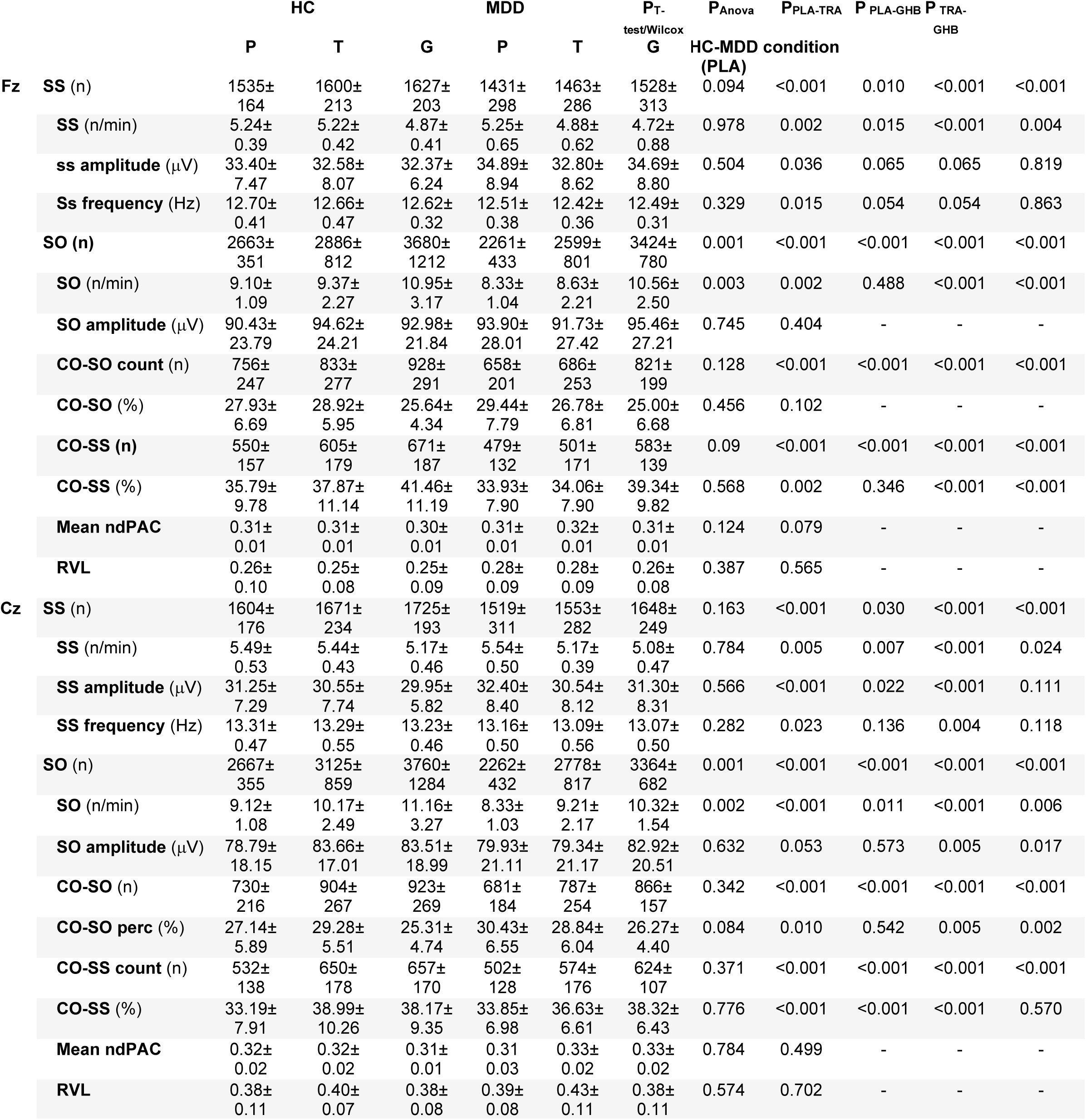
The table summarizes group differences and condition effects on spindle, slow oscillation, and spindle-slow oscillation coupling parameters during REM and NREM sleep, in Cz and Fz electrodes. Results are shown separately for healthy controls (HC) and patients with major depressive disorder (MDD). Between-group differences within each condition (PLA, TRA, GHB) were assessed using independent-samples t-tests or Wilcoxon rank-sum tests. Effects of condition are reported in the column “P_Anova_”. Benjamini-Hochberg corrected post-hoc pairwise comparisons between conditions were performed using estimated marginal means. Statistical significance was defined as p < 0.05. SS, Sleep spindles; SO, Slow oscillations; CO-SO, slow oscillations with co-occurring spindle; CO-SS, spindles with co-occurring slow oscillations; ndPAC, normalized direct phase-amplitude coupling; RVL, relative vector length.

|  |  | HC |  |  | MDD |  |  | P<sub>T-test/Wilcox</sub> | P<sub>Anova</sub> | P<sub>PLA-TRA</sub> | P<sub>PLA-GHB</sub> | P<sub>TRA-GHB</sub> |
|---|---|---|---|---|---|---|---|---|---|---|---|---|
|  |  | P | T | G | P | T | G | HC-MDD condition (PLA) |  |  |  |  |
| **Fz** | **SS** (n) | 1535±164 | 1600±213 | 1627±203 | 1431±298 | 1463±286 | 1528±313 | 0.094 | <0.001 | 0.010 | <0.001 | <0.001 |
|  | **SS** (n/min) | 5.24±0.39 | 5.22±0.42 | 4.87±0.41 | 5.25±0.65 | 4.88±0.62 | 4.72±0.88 | 0.978 | 0.002 | 0.015 | <0.001 | 0.004 |
|  | **ss amplitude** (µV) | 33.40±7.47 | 32.58±8.07 | 32.37±6.24 | 34.89±8.94 | 32.80±8.62 | 34.69±8.80 | 0.504 | 0.036 | 0.065 | 0.065 | 0.819 |
|  | **Ss frequency** (Hz) | 12.70±0.41 | 12.66±0.47 | 12.62±0.32 | 12.51±0.38 | 12.42±0.36 | 12.49±0.31 | 0.329 | 0.015 | 0.054 | 0.054 | 0.863 |
|  | **SO (n)** | 2663±351 | 2886±812 | 3680±1212 | 2261±433 | 2599±801 | 3424±780 | 0.001 | <0.001 | <0.001 | <0.001 | <0.001 |
|  | **SO** (n/min) | 9.10±1.09 | 9.37±2.27 | 10.95±3.17 | 8.33±1.04 | 8.63±2.21 | 10.56±2.50 | 0.003 | 0.002 | 0.488 | <0.001 | <0.001 |
|  | **SO amplitude** (µV) | 90.43±23.79 | 94.62±24.21 | 92.98±21.84 | 93.90±28.01 | 91.73±27.42 | 95.46±27.21 | 0.745 | 0.404 | - | - | - |
|  | **CO-SO count** (n) | 756±247 | 833±277 | 928±291 | 658±201 | 686±253 | 821±199 | 0.128 | <0.001 | <0.001 | <0.001 | <0.001 |
|  | **CO-SO** (%) | 27.93±6.69 | 28.92±5.95 | 25.64±4.34 | 29.44±7.79 | 26.78±6.81 | 25.00±6.68 | 0.456 | 0.102 | - | - | - |
|  | **CO-SS (n)** | 550±157 | 605±179 | 671±187 | 479±132 | 501±171 | 583±139 | 0.09 | <0.001 | <0.001 | <0.001 | <0.001 |
|  | **CO-SS** (%) | 35.79±9.78 | 37.87±11.14 | 41.46±11.19 | 33.93±7.90 | 34.06±7.90 | 39.34±9.82 | 0.568 | 0.002 | 0.346 | <0.001 | <0.001 |
|  | **Mean ndPAC** | 0.31±0.01 | 0.31±0.01 | 0.30±0.01 | 0.31±0.01 | 0.32±0.01 | 0.31±0.01 | 0.124 | 0.079 | - | - | - |
|  | **RVL** | 0.26±0.10 | 0.25±0.08 | 0.25±0.09 | 0.28±0.09 | 0.28±0.09 | 0.26±0.08 | 0.387 | 0.565 | - | - | - |
| **Cz** | **SS** (n) | 1604±176 | 1671±234 | 1725±193 | 1519±311 | 1553±282 | 1648±249 | 0.163 | <0.001 | 0.030 | <0.001 | <0.001 |
|  | **SS** (n/min) | 5.49±0.53 | 5.44±0.43 | 5.17±0.46 | 5.54±0.50 | 5.17±0.39 | 5.08±0.47 | 0.784 | 0.005 | 0.007 | <0.001 | 0.024 |
|  | **SS amplitude** (µV) | 31.25±7.29 | 30.55±7.74 | 29.95±5.82 | 32.40±8.40 | 30.54±8.12 | 31.30±8.31 | 0.566 | <0.001 | 0.022 | <0.001 | 0.111 |
|  | **SS frequency** (Hz) | 13.31±0.47 | 13.29±0.55 | 13.23±0.46 | 13.16±0.50 | 13.09±0.56 | 13.07±0.50 | 0.282 | 0.023 | 0.136 | 0.004 | 0.118 |
|  | **SO** (n) | 2667±355 | 3125±859 | 3760±1284 | 2262±432 | 2778±817 | 3364±682 | 0.001 | <0.001 | <0.001 | <0.001 | <0.001 |
|  | **SO** (n/min) | 9.12±1.08 | 10.17±2.49 | 11.16±3.27 | 8.33±1.03 | 9.21±2.17 | 10.32±1.54 | 0.002 | <0.001 | 0.011 | <0.001 | 0.006 |
|  | **SO amplitude** (µV) | 78.79±18.15 | 83.66±17.01 | 83.51±18.99 | 79.93±21.11 | 79.34±21.17 | 82.92±20.51 | 0.632 | 0.053 | 0.573 | 0.005 | 0.017 |
|  | **CO-SO** (n) | 730±216 | 904±267 | 923±269 | 681±184 | 787±254 | 866±157 | 0.342 | <0.001 | <0.001 | <0.001 | <0.001 |
|  | **CO-SO perc** (%) | 27.14±5.89 | 29.28±5.51 | 25.31±4.74 | 30.43±6.55 | 28.84±6.04 | 26.27±4.40 | 0.084 | 0.010 | 0.542 | 0.005 | 0.002 |
|  | **CO-SS count** (n) | 532±138 | 650±178 | 657±170 | 502±128 | 574±176 | 624±107 | 0.371 | <0.001 | <0.001 | <0.001 | <0.001 |
|  | **CO-SS** (%) | 33.19±7.91 | 38.99±10.26 | 38.17±9.35 | 33.85±6.98 | 36.63±6.61 | 38.32±6.43 | 0.776 | <0.001 | <0.001 | <0.001 | 0.570 |
|  | **Mean ndPAC** | 0.32±0.02 | 0.32±0.02 | 0.31±0.01 | 0.31 0.03 | 0.33±0.02 | 0.33±0.02 | 0.784 | 0.499 | - | - | - |
|  | **RVL** | 0.38±0.11 | 0.40±0.07 | 0.38±0.08 | 0.39±0.08 | 0.43±0.11 | 0.38±0.11 | 0.574 | 0.702 | - | - | - |

