## Supplementary Methods for "Targeting memory consolidation in major depression via pharmacological slow-wave sleep promotion"

### Pharmacological slow-wave sleep promotion does not enhance memory consolidation in major depression

#### Supplementary methods

***Inclusion and exclusion criteria.*** An experienced study physician assessed all participants regarding their general health and psychiatric history. Inclusion criteria for the major depressive disorder (MDD) group were: a diagnosis of MDD according to the *Diagnostic and Statistical Manual of Mental Disorders (DSM-5) [1],* stable antidepressant treatment (e.g., selective serotonin reuptake inhibitors [SSRIs] or serotonin–norepinephrine reuptake inhibitors [SNRIs]); age between 20 and 65 years; and no or low nicotine dependence, defined as a total score < 3 on the Fagerström Test for Nicotine Dependence [2].

Exclusion criteria for the MDD group entailed: known hypersensitivity or allergy to trazodone; daily intake of contraindicated drugs according to the Swiss federal medical product information (e.g., barbiturates, opioids); intake of any sleep-promoting medication, such as benzodiazepines or z-drugs three days before an experimental night or on a regular basis; any axis-I DSM-5 psychiatric disorder other than MDD (e.g., schizophrenia, bipolar disorder); neurological disorders or head injury; any clinically relevant medical diseases; family history of axis-I psychiatric disorders other than depression and stress-related disorders (e.g., schizophrenia or bipolar disorder); any form of substance dependence or regular use of illegal substances (lifetime use per substance greater than five times, with exception of occasional cannabis use); lifetime history of GHB use; for female participants: pregnancy, breast feeding, planned pregnancy during the course of the study, or lack of safe contraception (self-reported); inability to follow the procedures of the study because of language problems (i.e., attendance of primary school in a German-speaking country required); participation in another study with investigational psychoactive substances within 30 days preceding and during the present study; dietary restrictions. Prior to definitive enrollment into the study, all participants underwent a polysomnography (PSG) examination to exclude sleep-related disorders such as sleep apnea, restless legs syndrome and sleep onset REM sleep.

Exclusion criteria for the Healthy Control group were the same as in patients plus the presence of any axis-I DSM-5 psychiatric disorder.

On experimental days, participants were instructed to abstain from caffeine after 13:00h and from alcohol for at least 24 hours prior to measurements. On each experimental night, urine samples were taken to ensure abstinence from illegal substance use and to exclude pregnancy in female participants. A sleep-wake diary as well as a rest-activity monitor (GENEActiv, Activinsights Ltd., United Kingdom) worn on the wrist of the non-dominant hand were used to monitor the sleep-wake schedule.

**Detailed clinical assessments.**

Screening night: Structured interviews for depressive symptoms were conducted by an experienced study physician with at least 3 years of clinical psychiatry experience. Clinician-administered questionnaires included the Montgomery-Åsberg Depression Rating Scale (MADRS) [3] and the Hamilton Depression Rating Scale (HAMD) [4]. Self-reported sleep quality was evaluated during the screening session using the Pittsburgh Sleep Quality Index (PSQI) [5].

Experimental nights: Self-reported depressive symptoms were measured at the start of each session with the Beck Depression Inventory (BDI, 21:30) [6]. The Positive and Negative Affective Scale (PANAS) [7] was applied to assess momentary positive and negative affect before (22:30) and in the morning after (09:30) the experimental night (24:00-07:30). The Karolinska Sleepiness Scale (KSS) [8] was used to evaluate current sleepiness immediately before drug administration (23:30) and after awakening (07:35). An evening questionnaire (EQ, 23:30) was administered to record intake of caffeinated or alcoholic drinks, daytime naps, and medication use. After awakening, a Morning Questionnaire (MQ, 08:00) [9] was used to assess subjective sleep quality for each experimental night.

In the MQ, participants were asked to estimate time awake after lights-off (min), report the number of nocturnal awakenings (count) during the previous night, and rate sleep-related items on visual analogue scales (range 1-100) (sleep: "deep" vs. "superficial"; sleep: "quiet" vs. "restless"; current state: "recovered" vs. "tired"; current energy level: "full of energy" vs. "listless"). A suicide screening form (adapted from the suicide status form II) [10] was administered to MDD patients at the end of each session to rule out acute suicidality.

**Polysomnography data acquisition.** During the experimental nights, polysomnographic (PSG) recordings were conducted in accordance with the guidelines of the American Academy of Sleep Medicine (AASM) [11]. Recordings included electroencephalography (EEG), bipolar electrooculography (EOG), electromyography (EMG), and electrocardiography (ECG). For EEG acquisition, 23 electrodes were positioned according to the international 10–20 system.[12] Of these, 20 electrodes were used for EEG recording (Fp1, Fp2, F3, F4, F7, F8, Fz, T3, T4, T5, T6, C3, C4, Cz, P3, P4, Pz, O1, O2, Oz), two served as reference electrodes (A1, A2), and one functioned as a ground electrode placed on the forehead. Electrode placement was determined by measuring head circumference and marking the corresponding locations using a fitted cap (EasyCap GmbH, Herrsching, Germany). EOG electrodes were positioned diagonally across the eyes (E1, E2). Submental EMG activity was recorded using three electrodes placed on the chin (Chin1, Chin2, ChinZ), and two additional EMG electrodes were vertically attached over the tibialis posterior muscle of the right leg. Overnight PSG recordings were obtained using a dedicated PSG amplifier (SIENNA Ultimate, EMS Handels GmbH, Korneuburg, Austria). Signals were processed with a 50 Hz notch filter to eliminate line noise, along with high-pass filters (EEG: 0.5 Hz; EMG: 5 Hz; ECG: 1 Hz) and anti-aliasing low-pass filters (EEG: 70 Hz; EMG: 100 Hz; ECG: 70 Hz). Data were digitized and stored at a sampling rate of 512 Hz.

***Polysomnography data processing.*** Recordings were segmented into 30-second epochs and a sleep stage was determined for each epoch. Visual sleep stage scoring was done using Rembrandt® Analysis Manager (version 8; Embla Systems) according to the criteria of the AASM [11] by two independent scorers. Discrepancies across scorers were resolved by a third scoring expert. For sleep stage scoring, EEG derivations F3, C3, and O1, as well as EOG E1 and E2 were referenced to linked mastoids. For chin muscle tone, either Chin1 or Chin2 referenced against the central chin electrode (ChinZ) were considered. Study participants could choose their bedtime (between 22:00 to 24:00) but to simplify descriptions and data presentations, we will only refer to the 24:00–07:30 rhythm. As the study medications were administered at two points during the night, with trazodone or solid placebo at 23:30 and GHB or liquid placebo at 03:30, total time in bed (TIB) was considered as the time from lights-off to lights-on of the first (24:00-03:30) and second (03:30-07:30) night halves. The median time required for the administration of the liquid drug during the night from lights on to lights off was 3.5 min (range: 1 to 18 min) including an optional toilet break if requested. For the analysis of sleep architecture, sleep variables were computed for TIB. The following sleep variables were computed: (i) total sleep time (TST); (ii) sleep onset latency (SOL); (iii) wake after sleep onset (WASO); (iv) duration of sleep stages (i.e., non-REM [NREM] stage 1 [N1], NREM stage 2 [N2], NREM stage 3 [N3, SWS], and REM sleep); and (v) sleep efficiency ([SE] = [TST/TIB] × 100). The duration of sleep stages is reported as a percentage of TST.

***EEG preprocessing.*** EEG data were preprocessed based on the procedures described in Bavato et al. 2025 [13]. Data were analyzed using MATLAB (R2024a, MathWorks Inc.) and the EEGLAB toolbox (eeglab2023.1). In a first step, data were down-sampled to 128 Hz. EEG channels were bandpass filtered between 0.5-40 Hz and re-referenced to linked mastoids. Transient artifacts were detected as follows: First, bad EOG and EMG channels were flagged using a standard-deviation–based procedure on sleep-only samples (EOG: SD > 20; EMG: SD > 15). EMG movement artifacts were detected by applying a 25-90 Hz EMG bandpass filter, computing EMG band-power in 1-second windows and identifying epochs exceeding the 93rd percentile of each EMG channel's distribution. Artifact-labeled epochs were merged, padded ±4 s, and validated by a concurrent increase in ≥75% of canonical EEG channels (F3, F4, C3, C4, P3, P4, O1, O2) surpassing the 90th percentile of EEG band-power. EEG high-frequency artifacts were detected using a 30-90 Hz filter and a Hilbert envelope threshold defined per channel as median + 0.9×MAD, evaluated on sleep-only data. A segment was accepted as an artifact if ≥80% of EEG channels exceeded the channel threshold in ≥70% of the samples. Eye movements were detected within N1 and REM segments when EOG power in the 0.3-2 Hz range exceeded the median + 5×MAD, accompanied by an increase in EEG ROI power surpassing its median + 3×MAD, and a negative correlation (r < -0.2) between the two EOG channels. EEG Channels were visually inspected and flagged as bad in only the first half, only the second half, or the whole night. Corresponding sections of affected channels were interpolated using spherical interpolation (EEGLAB's *pop_interp*). Cardiac (R-peak) artifacts were removed as done in Stucky et al 2026

[14], with wake periods and artefactual time periods excluded from the fitting process. Electrical noise was filtered by applying 50 Hz and 100 Hz notch filters and an FFT-based narrowband noise reduction method targeting device-specific peaks (multiples of 9.1 Hz). At affected frequencies, amplitude was replaced with the 1 Hz moving median of neighboring frequencies, followed by phase correction and inverse FFT reconstruction of the cleaned signal.

***Quantitative EEG analysis.*** For quantitative EEG analysis, the data were divided into 30-second epochs, and only segments free of artifacts were retained for analysis. For each epoch, power spectral density (PSD) was calculated using Welch's method with 4-second Hanning windows, 50% overlap, and a frequency resolution of 0.5 Hz. PSD values were derived from the C3 electrode across a frequency range of 0.5 to 20 Hz, and power values were expressed in decibels ($10 \cdot \log_{10}(\mu V^2/Hz)$).

Spectral power bands of the EEG were defined as follows: delta 1 (d1, 0.5-1 Hz), delta 2 (d2, 2-4 Hz), theta 1 (t1, 4-6 Hz), theta 2 (t2, 6-8 Hz), alpha 1 (a1, 8-10 Hz), alpha 2 (a2, 10-12 Hz), sigma 1 (s1, 11-13 Hz), sigma 2 (s2, 13-16 Hz), beta 1 (b1, 15-20 Hz), beta 2 (b2, 20-30 Hz).

***Spindle detection***. Following filtering and cleaning of the data, sleep spindles were automatically detected using the *yasa.spindles_detect* function in the python-based YASA algorithm (Version 0.6.5) using Matlab's built-in support for Python integration. Spindles were detected in the 11-16 Hz band in Fz and Cz EEG channels [15]. For the analysis, only spindles <100μV in amplitude (see Staresina et al, 2015) were included to exclude artefacts. Fixed bandwidths were used to separate slow spindles on Fz (11-13 Hz) and fast spindles on Cz (13-16 Hz), matching their frontal, respectively central, predominance in the EEG [16]. Root-mean-square (RMS) values exceeding the individual threshold for 0.5 to 3 seconds were identified as spindles. Similar to Barbaux et al 2025, the RMS threshold was set to 2.5 SD and quality of the spindle detection algorithm was verified by manual inspection. Detected spindles closer than 500ms were considered the same spindle. Spindle densities were calculated by dividing spindle counts by the minutes spent in NREM2+3 stages for each individual, condition and channel. Detected spindle densities were within normal ranges.[16]

***Slow-wave detection**.* Slow-waves were automatically detected in Fz and Cz EEG channels from separate and artefact-free N2 and N3 sleep stages using the *yasa.sw_detect* function of the YASA algorithm (Version 0.6.5) [17]. Slow-waves were detected for frequencies between 0.5-1.25 Hz. Only strong slow oscillations, defined as PTP values above the participant- and channel-specific 75$^{th}$ percentile during the placebo condition, were retained for coupling analyses.

***Slow-oscillation – spindle co-occurrence and coupling.*** The following coupling metrics of slow-oscillations and spindles were derived from the *yasa.sw_detect* and *yasa.spindles_detect* functions and analyzed for concatenated N2/N3 stages:

Co-occurrence of discrete events. Slow-waves and spindles were considered co-occurring (SS-SO) if the spindle amplitude peak occurred within ±1.2s of the negative slow-oscillation peak [15]. Also, the reverse case was calculated, where the number of slow oscillations happening within ±1.2s of the spindle peak (SO-SS) were calculated. For each participant, condition, and channel, the proportion of co-occurring spindles and slow oscillations was calculated relative to the total number of detected slow-oscillation events above the 75$^{th}$ amplitude percentile. The density of co-occurring events was calculated by dividing co-occurrence counts by the minutes spent in NREM2+3 stages for each individual, condition and channel.

Coupling phase. The coupling phase was calculated relative to the negative slow wave peak, within a window of ±2s. For this, sigma peak phase was calculated as the timepoint where maximal sigma-filtered (11 - 16 Hz) amplitude occurred relative to the slow oscillation cycle. Phase at sigma peak thus refers to the time point where sigma peak occurred on the slow-wave filtered signal (0.5-1.5 Hz) in radians, and was reported averaged across co-occurring events for each participant, condition, and channel.

The consistency of phase alignment across events was additionally quantified using the resultant vector length (RVL), with group differences in preferred phase assessed using circular statistics. The phase of the slow oscillation at the spindle peak was obtained from the slow-wave filtered signal (0.5-1.5 Hz) using a Hilbert transform, as implemented in the YASA detection pipeline. RVL was computed across all coupling events per participant, condition, and channel. A longer RVL suggests that most spindles are closely clustered around one mean slow oscillation phase, indicating stronger spindle-slow oscillation coupling. A short RVL indicates greater variance of spindles on the slow oscillation phase, indicating weaker coupling.

Coupling strength. Normalized direct phase–amplitude coupling (ndPAC), a variant of the mean vector length approach, was used to quantify the coupling strength between instantaneous slow-oscillation phase and sigma-band amplitude. ndPAC was computed for each slow oscillation using the yasa.sw_detect function [17] [18]. In contrast to the classical mean vector length metric, ndPAC includes z-scoring of the amplitude signal to reduce bias from direct current components and applies an internal statistical threshold to suppress spurious coupling estimates, thereby removing the need for surrogate-based normalization. Coupling values range from 0 (no coupling) to 1 (perfect coupling). The ndPAC value was calculated relative to the negative slow-wave peak within a ±2 s window. Only values greater than zero and restricted to slow oscillations with a detected co-occurring spindle event were included to derive mean ndPAC per participant, condition, and channel.

#### Supplementary results

##### Declarative, procedural and emotional memory

| Variable | Time | Group | PLA | TRA | GHB |
|---|---|---|---|---|---|
| **Cycles (n)** | Evening | HC | 21.57 ± 4.69 | 20.98 ± 4.56 | 21.67 ± 4.05 |
| | Evening | MDD | 19.62 ± 4.99 | 18.65 ± 4.99 | 19.69 ± 4.83 |
| | Morning | HC | 26.09 ± 5.26 | 24.21 ± 5.02 | 25.65 ± 4.47 |
| | Morning | MDD | 23.52 ± 6.35 | 22.25 ± 6.60 | 24.00 ± 6.43 |
| **Accuracy (%)** | Evening | HC | 95.30 ± 2.81 | 94.76 ± 2.73 | 95.15 ± 2.51 |
| | Evening | MDD | 95.05 ± 3.73 | 95.50 ± 2.67 | 94.10 ± 3.63 |
| | Morning | HC | 96.39 ± 2.52 | 95.12 ± 5.06 | 94.35 ± 4.67 |
| | Morning | MDD | 95.92 ± 3.05 | 96.31 ± 3.13 | 96.77 ± 3.83 |

| | | | | | |
|---|---|---|---|---|---|
| **Mean reaction time (ms)** | Evening | HC | 269.86 ± 76.18 | 277.82 ± 80.12 | 261.86 ± 51.98 |
| | Evening | MDD | 309.13 ± 108.85 | 327.48 ± 108.89 | 307.62 ± 109.45 |
| | Morning | HC | 205.00 ± 49.09 | 227.68 ± 66.96 | 206.67 ± 41.41 |
| | Morning | MDD | 247.09 ± 98.71 | 271.70 ± 115.87 | 242.62 ± 105.09 |
| **Median reaction time (ms)** | Evening | HC | 233.90 ± 60.58 | 238.50 ± 64.07 | 224.67 ± 44.05 |
| | Evening | MDD | 267.13 ± 95.93 | 282.33 ± 91.81 | 266.67 ± 97.14 |
| | Morning | HC | 188.52 ± 46.75 | 206.86 ± 55.87 | 183.52 ± 39.57 |
| | Morning | MDD | 215.22 ± 85.75 | 240.78 ± 100.15 | 212.00 ± 92.30 |
| **Mean speed (1/s)** | Evening | HC | 5.76 ± 1.88 | 6.38 ± 2.48 | 6.73 ± 2.48 |
| | Evening | MDD | 6.54 ± 3.80 | 5.11 ± 2.32 | 6.17 ± 2.66 |
| | Morning | HC | 7.39 ± 1.90 | 8.33 ± 4.51 | 9.41 ± 3.47 |
| | Morning | MDD | 8.37 ± 6.03 | 6.49 ± 3.03 | 7.68 ± 4.24 |
| **Median speed (1/s)** | Evening | HC | 4.75 ± 1.35 | 4.63 ± 1.20 | 4.83 ± 1.15 |
| | Evening | MDD | 4.28 ± 1.31 | 4.01 ± 1.26 | 4.30 ± 1.38 |
| | Morning | HC | 5.70 ± 1.67 | 5.23 ± 1.46 | 5.77 ± 1.41 |
| | Morning | MDD | 5.34 ± 1.89 | 4.82 ± 1.77 | 5.46 ± 1.95 |

**Supplementary Table S3.** The table summarizes descriptive outcomes of the finger-tapping task (FTT) across experimental conditions (PLA, TRA, GHB), shown separately for healthy controls (HC) and patients with major depressive disorder (MDD), and for evening and morning assessments. Data are presented as mean ± standard deviation (SD). PLA, placebo; TRA, trazodone; GHB, sodium oxybate; FTT, finger-tapping task.

| Variable | Time | Group | PLA | TRA | GHB |
|---|---|---|---|---|---|
| **Accuracy (%)** | Evening | HC | 37.11 ± 18.81 | 43.42 ± 18.86 | 41.97 ± 20.91 |
| | Evening | MDD | 37.86 ± 15.56 | 38.92 ± 13.69 | 39.52 ± 14.65 |
| | Morning | HC | 63.29 ± 17.32 | 68.16 ± 17.77 | 65.66 ± 19.51 |
| | Morning | MDD | 56.43 ± 20.59 | 60.66 ± 16.18 | 56.67 ± 20.73 |

| | | | | | |
|---|---|---|---|---|---|
| **Accuracy (n)** | Evening | HC | 14.84 ± 7.52 | 17.37 ± 7.54 | 16.79 ± 8.36 |
| | Evening | MDD | 15.14 ± 6.22 | 15.55 ± 5.45 | 15.81 ± 5.86 |
| | Morning | HC | 25.32 ± 6.93 | 27.26 ± 7.11 | 26.26 ± 7.80 |
| | Morning | MDD | 22.57 ± 8.24 | 24.23 ± 6.44 | 22.67 ± 8.29 |
| **Learned words (n)** | Morning | HC | 11.74 ± 4.21 | 11.11 ± 3.36 | 11.21 ± 5.28 |
| | Morning | MDD | 9.19 ± 3.91 | 10.14 ± 4.39 | 9.24 ± 4.37 |
| **Forgotten words (n)** | Morning | HC | 1.26 ± 1.10 | 1.21 ± 1.08 | 1.74 ± 1.37 |
| | Morning | MDD | 1.76 ± 1.67 | 1.50 ± 1.44 | 2.38 ± 3.63 |
| **Remembered words (n)** | Morning | HC | 13.58 ± 7.60 | 16.16 ± 7.60 | 15.05 ± 8.10 |
| | Morning | MDD | 13.38 ± 6.24 | 14.05 ± 5.55 | 13.43 ± 6.34 |

**Supplementary Table S4.** The table summarizes descriptive outcomes of the paired-word learning task (PWLT) across experimental conditions (PLA, TRA, GHB), shown separately for healthy controls (HC) and patients with major depressive disorder (MDD), and for evening and morning assessments. Data are presented as mean ± standard deviation (SD). ‘Learned words’ refer to word pairs recalled correctly only in the morning, ‘forgotten words’ to word pairs recalled correctly only in the evening, and ‘remembered words’ to word pairs recalled correctly at both assessments. PLA, placebo; TRA, trazodone; GHB, sodium oxybate; PWLT, paired-word learning task.

227

| **Variable** | **Group** | **PLA** | **TRA** | **GHB** |
|---|---|---|---|---|
| **Positive correct (proportion)** | HC | 0.61 ± 0.19 | 0.60 ± 0.19 | 0.60 ± 0.15 |
| | MDD | 0.63 ± 0.20 | 0.62 ± 0.21 | 0.63 ± 0.21 |
| **Neutral correct (proportion)** | HC | 0.54 ± 0.27 | 0.47 ± 0.18 | 0.52 ± 0.18 |
| | MDD | 0.56 ± 0.21 | 0.61 ± 0.25 | 0.55 ± 0.25 |
| **Negative correct (proportion)** | HC | 0.75 ± 0.15 | 0.71 ± 0.18 | 0.70 ± 0.15 |
| | MDD | 0.71 ± 0.16 | 0.72 ± 0.23 | 0.74 ± 0.14 |
| **True positive (proportion)** | HC | 0.30 ± 0.09 | 0.29 ± 0.08 | 0.28 ± 0.07 |
| | MDD | 0.30 ± 0.08 | 0.30 ± 0.10 | 0.30 ± 0.09 |

| | | | | |
|---|---|---|---|---|
| **True negative (proportion)** | HC | 0.46 ± 0.08 | 0.47 ± 0.03 | 0.47 ± 0.04 |
| | MDD | 0.44 ± 0.09 | 0.43 ± 0.10 | 0.45 ± 0.09 |
| **False negative (proportion)** | HC | 0.20 ± 0.10 | 0.21 ± 0.08 | 0.20 ± 0.07 |
| | MDD | 0.21 ± 0.10 | 0.22 ± 0.13 | 0.20 ± 0.09 |
| **True total (proportion)** | HC | 0.75 ± 0.10 | 0.75 ± 0.07 | 0.75 ± 0.06 |
| | MDD | 0.74 ± 0.09 | 0.73 ± 0.12 | 0.75 ± 0.10 |
| **False total (proportion)** | HC | 0.25 ± 0.10 | 0.25 ± 0.07 | 0.25 ± 0.06 |
| | MDD | 0.26 ± 0.09 | 0.27 ± 0.12 | 0.25 ± 0.10 |
| **d' (a.u.)** | HC | 1.66 ± 0.56 | 1.70 ± 0.38 | 1.68 ± 0.43 |
| | MDD | 1.51 ± 0.50 | 1.36 ± 1.00 | 1.48 ± 0.91 |

**Supplementary Table S5.** The table summarizes descriptive outcomes of the emotional picture memory task (EPMT) across experimental conditions (PLA, TRA, GHB), shown separately for healthy controls (HC) and patients with major depressive disorder (MDD). Data are presented as mean ± standard deviation (SD). Recognition outcomes are reported as proportions. Recognition sensitivity (d′) was calculated as Z(hit rate) − Z(false-alarm rate). PLA, placebo; TRA, trazodone; GHB, sodium oxybate; EPMT, emotional picture memory task; d′, recognition sensitivity.

#### Statistics: Group – condition interactions

***EEG polysomnographic outcomes.*** Significant group-condition interactions were observed for PLA–TRA for WASO ($p_{FDR}$ = 0.032, ES = 1.20) and SE ($p_{FDR}$ = 0.043, ES = −1.02), and for PLA–GHB for SE ($p_{FDR}$ = 0.042, ES = −1.16) and TST ($p_{FDR}$ = 0.046, ES = −1.07), indicating stronger sleep promoting effects of both drugs in MDD than HC.

***EEG spectral outcomes.*** Significant group-condition interactions during REM sleep were observed for PLA-GHB for the aperiodic exponent ($p_{FDR}$ = 0.017, ES = −1.31), aperiodic offset ($p_{FDR}$ = 0.021, ES = −1.22), relative alpha ($p_{FDR}$ = 0.049, ES = 1.13), beta2 ($p_{FDR}$ = 0.032, ES = 1.21), and beta power ($p_{FDR}$ = 0.034, ES = 1.20). For TRA-GHB, significant group-condition interactions were observed for the aperiodic exponent ($p_{FDR}$ = 0.023, ES = −1.15) and aperiodic offset ($p_{FDR}$ = 0.021, ES = −1.16). During NREM sleep, significant group-condition interactions (MDD-HC) were observed for PLA-GHB for relative delta2 power ($p_{FDR} = 0.026$, ES = 1.12) and for TRA-GHB for relative delta2 power ($p_{FDR} = 0.014$, ES = 1.33).

**EEG spindle-slow oscillation temporal and phase coupling.**

Spindle–slow oscillation coupling: At Fz, group-by-condition interactions were restricted to slow-oscillation count (PLA-TRA and PLA-GHB: both $p_{FDR} < 0.001$; TRA-GHB: $p_{FDR} = 0.021$). No other drug effects or interactions were significant after FDR correction. At Cz, Group-by-condition interactions were observed for spindle count, slow-oscillation count, and co-occurring event counts (all $p_{FDR} \leq 0.035$); no other drug effects or interactions were significant after FDR correction.

Preferred phase: A group-by-condition interaction was observed only for TRA-GHB at Cz ($p_{FDR} = 0.039$; −11.79°). No other preferred-phase interactions were significant after FDR correction.

Phase-based spindle–slow oscillation coupling: A significant group-condition interaction (MDD-HC) was observed at Cz for TRA-GHB preferred spindle phase relative to the slow-oscillation cycle ($p_{FDR} = .039$, phase difference = −11.79°), indicating a larger TRA-GHB phase shift in MDD than HC. No significant group-condition interactions (MDD-HC) were observed for preferred spindle phase at Fz (all $p_{FDR} \geq .846$), or for PLA-TRA and PLA-GHB at Cz ($p_{FDR} = .814$ and .161, respectively).
